# Performance Evaluation of the Verily Numetric Watch sleep suite for digital sleep assessment against in-lab polysomnography

**DOI:** 10.1101/2024.09.10.24313425

**Authors:** Benjamin W. Nelson, Sohrab Saeb, Poulami Barman, Nishant Verma, Hannah Allen, Massimiliano de Zambotti, Fiona C. Baker, Nicole Arra, Niranjan Sridhar, Shannon S. Sullivan, Scooter Plowman, Erin Rainaldi, Ritu Kapur, Sooyoon Shin

## Abstract

The goal was to evaluate the performance of a multi-sensor wrist-worn wearable device for generating 12 sleep measures in a diverse cohort. Our study technology was the sleep suite of the Verily Numetric Watch (VNW), using polysomnography (PSG) as reference during 1-night simultaneous recording in a sample of N=41 (18 male, age range: 18-78 years). We performed epoch-by-epoch comparisons for all measures. Key specific analyses were: core accuracy metrics for sleep vs wake classification; bias for continuous measures (Bland-Altman); Cohen’s kappa and accuracy for sleep stage classifications; and mean count difference and linearly weighted Cohen’s kappa for count metric. In addition, we performed subgroup analyses by sex, age, skin tone, body mass index, and arm hair density. Sensitivity and specificity (95% CI) of sleep versus wake classification were 0.97 (0.96, 0.98) and 0.66 (0.61, 0.71), respectively. Mean total sleep time bias was 14.55 minutes (1.61, 27.16); wake after sleep onset, −11.77 minutes (−23.89, 1.09); sleep efficiency, 3.15% (0.68, 5.57); sleep onset latency, −3.24 minutes (−9.38, 3.57); light-sleep duration, 3.78 minutes (−7.04, 15.06); deep-sleep duration, 3.91 minutes (−4.59, 12.60); rapid eye movement-sleep duration, 6.94 minutes (0.57, 13.04). Median difference for number of awakenings, 0.00 (0.00, 1.00); and overall accuracy of sleep stage classification, 0.78 (0.51, 0.88). Most measures showed statistically significant proportional biases and/or heteroscedasticity. Subgroup results appeared largely consistent with the overall group, although small samples preclude strong conclusions. These results support the use of VNW’s in classifying sleep versus wake, sleep stages, and for related overnight sleep measures.

## Introduction

Sleep behaviors may provide important insights into both mental and physical health status, as well as therapeutic and disease outcomes. Sleep disturbance constitutes a meaningful aspect of health, and inadequate sleep is associated with increased risk for major depressive disorder, cardiovascular disease, cancer, and diabetes.^1,2^

However, the traditional reference standard for measuring sleep staging, polysomnography (PSG), is resource-intensive and in limited supply. Measuring sleep via PSG imparts high burden and cost, and for many purposes, may diminish ecological validity.^3^ The emergence of user-friendly, low-burden, yet highly accurate and reliable digital technologies is a promising development. These technologies may open the door to long-term sleep monitoring, allowing for evaluation of changes in sleep over time (e.g., regularity, variance patterns, and trends over days, weeks, or months). Such methods may also facilitate the wider evaluation of sleep-related outcomes across multiple diseases where they remain under-studied despite their importance.

Wearable sensor-based sleep-tracking technology may lead to important advances towards these goals by providing accessible, convenient, long-term, free-living sleep behavior monitoring.^4^ However, the responsible deployment of these devices demands proper performance evaluation studies to ensure their accuracy and generalizability across demographic groups, ^5–7^ and to mitigate any potential biases that could lead to health disparities.^8–15^

Following the widespread adoption of sleep-tracking technology by the general public and the subsequent emergence of reports in the clinical literature,^16–18^ device performance evaluation studies have proliferated in recent years,^5–7^ although concerns remain about adoption for diagnosis and treatment.^19–21^ Those studies have shown that, in general, latest-generation wearable devices produce suitable estimates of sleep measures, although these vary by device and firmware version and their capabilities to classify sleep stages are less sufficient.^22,23^ Despite growing research to understand overall device performance, investigations about generalizability across diverse populations, including sex, age, skin tone, body mass index (BMI), and arm hair density are still lacking.^4^

We undertook this study in order to evaluate the analytic performance of sleep tracking by the Verily Numetric Watch (VNW), a wrist-worn device that classifies every 30-second epoch into one of the following 4 sleep stages: wake, light sleep, deep sleep, and rapid eye movement (REM) sleep, allowing for the derivation of a number of clinically meaningful overnight sleep measures including total sleep time (TST); wake after sleep onset (WASO); sleep efficiency (SE); sleep onset latency (SOL); number of awakenings (NAWK); and light, deep, and REM sleep duration.

Our objectives were to evaluate the VNW’s performance for epoch-by-epoch sleep versus wake classification; 4-stage sleep classification (wake, light, deep, REM); and TST, WASO, SE, SOL, NAWK, and sleep stage duration in a diverse sample of sleepers, compared to PSG-derived labels. In addition, the exploratory objectives included performing subgroup analyses based on sex, age, skin tone, BMI, and arm hair density.

## Methods

### Sample

This study included 41 participants. Eligible participants were between 18-80 years of age and agreed to abstain from caffeine, nicotine, alcohol, and cannabis products for eight hours prior to the overnight lab visit and until the visit was complete. They also had to agree to abstain from medications that may affect sleep/wakefulness for 24 hours prior to the lab visit and during the study, unless the medications were taken on a routine basis and had approval from the study team. In order to be eligible, participants had to be considered “typical sleepers,” based on the following: obstructive sleep apnea 50 (OSA-50)^24^ scores < 5; insomnia severity index (ISI)^25^ < 8; Epworth Sleepiness Scale (ESS) scores < 10; no evidence of sleep-disordered breathing at the PSG evaluation; and apnea-hypopnea index (AHI)^26^ threshold of 5 defining hypopnea as ≥ 30% reduction in airflow for ≥ 10 seconds associated with a ≥ 3% decrease in oxygen saturation or an arousal).^27^ Individuals were ineligible if they had a major medical or psychiatric condition, if they used supplemental oxygen, or were unwilling to cease use of therapy, such as continuous positive airway pressure or oral appliance for sleep-disordered breathing during the visit.

Additional exclusion criteria included use of medications that affect sleep (e.g., hypnotics or antidepressants) or any sleep medications in the previous 24 hours; pregnancy, lactation, or breastfeeding; having an implantable medical device; night-shift work; or travel over 3 time zones within two weeks prior to the study. The study was approved by the WCG Institutional Review Board (20215892) and was conducted in accordance with the Declaration of Helsinki. All participants provided informed consent, and the study was registered at clinicaltrials.gov (NCT05276362).

### Focus method/technology: Verily Numetric Watch

Participants wore the VNW wrist-worn device on the non-dominant wrist and this device was equipped with two sensors: a photoplethysmography (PPG) sensor with a sampling rate of 60 Hz and a 3-axis accelerometer with a sampling rate of 104 Hz. The PPG sensor consists of a green light emitter diode and two PPG signal channels. Using the PPG and accelerometer signals, the VNW classifies every 30-second epoch into one of the following 4 classes: wake, light sleep, deep sleep, and REM sleep.

The sleep staging algorithm consists of a deep convolutional neural network that was initially trained using 10,000 nights of data from the Sleep Heart Health Study (SHHS) and Multi-Ethnic Study of Atherosclerosis (MESA) public datasets,^28^ and tuned on a previous generation of the VNW with a dataset collected at SRI.

### Reference Labels

Standard laboratory PSG sleep assessment including electroencephalography (EEG; F3/4, C3/4, O1/2 referred to the contralateral mastoid; 256 Hz sampled), submental electromyography and bilateral electrooculography was performed according to the American Academy of Sleep Medicine (AASM) guidelines.^29^ Leg movement (bilateral anterior tibialis), electrocardiography (ECG), respiratory (thoracic and abdominal piezoelectric bands, nasal cannula and thermistor), and oxygen saturation (pulse oximeter) signals were also collected and used to confirm the absence of sleep disordered breathing. All recordings were performed using the Compumedics Grael® HD-PSG system (Compumedics, Abbotsford, Victoria, Australia). Sleep scoring (wake, N1, N2, N3, REM) was performed by two independent scorers according to the AASM rules. Inter-rater reliability (Kappa) was 91%. Discrepancies were resolved by a third scorer. To make the labels consistent with the VNW sleep suite, categories N1 and N2 light sleep were combined into a single “light sleep” category.^19^

### Design, study setting, and procedures

This was a single-arm, observational study to evaluate the performance of the sleep measures from the VNW against PSG-derived labels (as reference). Data were collected during a single overnight stay in a sleep laboratory from a diverse sample of sleepers without elevated insomnia symptoms or OSA. All study protocols and procedures were conducted at a single site (SRI; Menlo Park, California).

### Recruitment and Phone Screen

Potential participants were recruited through fliers, an existing site participant database, and postings on public websites. Study personnel pre-screened potential participants for eligibility, via phone and online screen questionnaires.

### In-Lab/Remote Screening and Enrollment Visit

After checking verbal interest and eligibility during a phone pre-screening, participants were invited to an in-lab or remote screening visit to sign an informed consent. Study personnel collected demographic, clinical, and other relevant information including skin tone and arm hair density. Candidates whose eligibility was confirmed in the in-lab/remote screening visit were scheduled for the in-lab overnight visit. Screen failures (defined as those who consented to participate but did not meet one or more eligibility criteria) were not entered in the study.

The questionnaires completed by participants or study personnel and used for screening were OSA50,^24^ AHI,^26^ ISI,^25^ ESS,^30^ Fitzpatrick Skin Scale,^31^ and Arm Hair Index (see Supplement).

### In-Lab Overnight Visit

Participants slept in comfortable sound-proof and temperature-controlled bedrooms where they were able to go to bed and wake up at their preferred times. During this visit, standard PSG protocols were used for preparation, recording procedures, and instrument calibration; and participants were outfitted with the VNW on their non-dominant wrist (see Supplement for additional information).

### Statistical Analysis

All analyses were conducted between lights off and lights on and structured to evaluate the performance of the VNW against the PSG reference, following published scientific recommendations for performance evaluation.^7^ The unit of analysis was the 30-second epoch. To evaluate the endpoints, all epochs with data from PSG and VNW from lights-out to lights-on were included in analyses.

Our core analysis included evaluation of sensitivity, specificity, negative predictive value (NPV) and positive predictive value (PPV) of sleep versus wake classification. Estimates of sensitivity, specificity, PPV, and NPV were obtained using all epochs that were non-missing in both devices. We accounted for clustering of epochs within an individual using logistic mixed-effect regression models, with subject added as random effect. 95% CIs were calculated using clustered bootstrap method.^32^ Additionally, we evaluated classification of different sleep stages (light, deep, REM) and derived sleep measures TST, WASO, SE, SOL, NAWK, and duration of different sleep stages (light, deep, REM) as part of the core analysis.

VNW’s performance for derived sleep measurements, at each participant level, were calculated using all epochs from lights-off to lights-on, based on existing performance testing standardization frameworks.^19,33^ For these measurements, we performed Bland Altman analyses, estimating the mean bias and lower and upper limits of agreement (with their 95% CIs), testing for assumptions of proportional bias, heteroscedasticity, and normality. 95% percentile bootstrap CIs are reported for TST, WASO, SE, SOL, NAWK, sleep stage duration. We evaluated the VNW’s measurements of NAWKs, using mean and median difference in counts, and linearly weighted Cohen’s kappa (with their 95% CIs). To assess the accuracy of 4-stage classification of sleep stages (light, deep, REM and wake) we report confusion matrix, overall Cohen’s kappa and accuracy, with associated 95% CIs.

Additionally for each sleep stage we report stage-specific Cohen’s Kappa, stage-specific accuracy, PPV and sensitivity. To obtain accuracy measures on each sleep stage, the outcomes were dichotomized to the sleep stage of interest against all others. The average method calculates the measure for each individual participant and then averages out the measure across all participants [19]. 95% percentile CIs were obtained using bootstrap method. All analyses were performed with R version 4.3.1 (2023-06-16).

As part of exploratory analysis, we further evaluated all core analytics endpoints across relevant participant subgroups (i.e., age, sex, BMI, skin tone, arm hair density). Analyses were performed among subgroups with a sample size ≥ 10. In some cases, groups with fewer than 10 participants were combined with other groups that shared similar characteristics, when possible, to obtain a minimum sample size of at least 10 participants.

## Results

Participants included 41 adults (18 male) with a mean age of 40.5 years (SD, 16.5) and ranging from 18-78 years. The majority of participants were White (22 [53.7%]) and not hispanic or latino (37 [90.2%]), but there was diversity in subgroups, including different skin tones (light, n=21; medium, n=15; dark, n=5), body mass index (BMI; range: 17.8 - 36.0), and arm hair density (little to no visible hair, n=17; visible fine hair, n=16; coarse and very coarse hair, n=8) (Table 1, Supplemental Table 1).

**Table 1.** Summary of key participant characteristics.

|  |  | **N=41** |
| --- | --- | --- |
| **Age (years)** | Median (range) | 34.0 (18.0 - 78.0) |
|  | Mean (SD) | 40.5 (16.5) |
| **Age categories, n (%)** | 18-40 | 25 (61.0) |
|  | 41-80 | 16 (39.02) |
| **Sex, n (%)** | Female | 23 (56.1) |
|  | Male | 18 (43.9) |
| **BMI categories, n (%)** | < 25 | 30 (73.17) |
|  | ≥ 25 | 11 (26.82) |
| **Skin tone, n (%)** | Light Skin Tone | 21 (51.21) |
|  | Medium Skin Tone | 15 (36.59) |
|  | Dark Skin Tone | 5 (12.20) |
| **Arm hair index, n (%)** | 1: Little to no visible arm hair, light in color | 17 (41.5) |
|  | 2: Visible, fine, arm hair, light to medium color | 16 (39.0) |
|  | 3 and 4: Coarse and very coarse arm hair, medium to dark color | 8 (19.51) |
| **Race, n (%)** | American Indian or Alaska Native | 1 (2.4) |
|  | Asian | 8 (19.5) |
|  | Black or African American | 4 (9.8) |
|  | Mixed race | 4 (9.8) |
|  | Native Hawaiian or Other Pacific Islander | 1 (2.4) |
|  | Other | 1 (2.4) |
|  | White | 22 (53.7) |
| **Ethnicity, n(%)** | Hispanic or Latino | 4 (9.76) |
|  | Not Hispanic or Latino | 37 (90.20) |
| **Dominant hand, n (%)** | Ambidextrous | 1 (2.4) |
|  | Left | 5 (12.2) |
|  | Right | 35 (85.4) |
| **OSA score** | Median (range) | 0.0 (0.0 - 7.0) |
|  | Mean (SD) | 1.3 (1.6) |
| **ISI score** | Median (range) | 3.0 (0.0 - 7.0) |
|  | Mean (SD) | 3.0 (1.9) |
| **ESS score** | Median (range) | 5.0 (0.0 - 9.0) |
|  | Mean (SD) | 5.0 (2.7) |
| **AHI Index** | Median (range) | 1.3 (0.1 - 4.7) |
|  | Mean (SD) | 1.7 (1.2) |

Note: AHI=Apnea hypopnea index ; BMI=body mass index; ESS=Epworth sleepiness scale ; ISI=insomnia severity index ; OSA=Obstructive Sleep Apnea; SD=standard deviation

### Core analytics and main outcome variables

We collected data for a total of 38,796 epochs and all epochs were used for analyses. The sensitivity of the VNW classifying sleep vs wake compared to the PSG was 0.96 (95% CI: 0.95, 0.98), specificity was 0.65 (95% CI: 0.60, 0.70), PPV was 0.92 (95% CI: 0.90, 0.94) and NPV was 0.79 (95% CI: 0.72, 0.88) (Table 2).

**Table 2.** Summary of performance metrics for ‘sleep vs wake’ classification of 30-second epochs.

|  | **Sensitivity [95% CI]** | **Specificity [95% CI]** | **NPV [95% CI]** | **PPV [95% CI]** |
| --- | --- | --- | --- | --- |
| **Sleep vs Wake** | 0.96 (0.95, 0.98) | 0.65 (0.60, 0.70) | 0.79 (0.72, 0.88) | 0.92 (0.90, 0.94) |

CI = confidence interval; NPV = negative predictive value (prediction of wake); PPV = positive predictive value (prediction of sleep)

The mean bias for TST was 14.55 minutes (95% CI: 1.61, 27.16), and for WASO was −11.77 minutes (−23.89, 1.09). For SE, the mean bias was 3.15% (95% CI: 0.68, 5.67) and for SOL, the mean bias was −3.24 minutes (95% CI: −9.38, 3.57). The mean bias for the duration of different sleep stages was 3.78 minutes (95% CI: −7.04, 15.06) for light sleep, 3.91 minutes (95% CI: −4.59, 12.60) for deep sleep, and 6.94 minutes (0.57, 13.04) for REM sleep. The median NAWK counts for PSG is 1 and VNW is 1 with the difference in median counts of NAWKs was 0.00 (95% CI: 0.00, 1.00), the difference in the mean counts of NAWKs was −0.02: (95% CI: −0.56, −0.54), and the linear weighted Cohen’s kappa coefficient was 0.41 (95% CI: 0.26, 0.57). Bland-Altman analyses showed that all measures had significant proportional bias (Table 3), with the VNW slightly overestimating values at the low end of the distribution, and underestimating them at the high end (Figure 1). For all measures, proportional bias was true; the assumption of normality was false for all measures, except for deep and REM sleep duration; and heteroscedasticity was false for all measures, except for SOL (Table 3).

**Figure 1.**
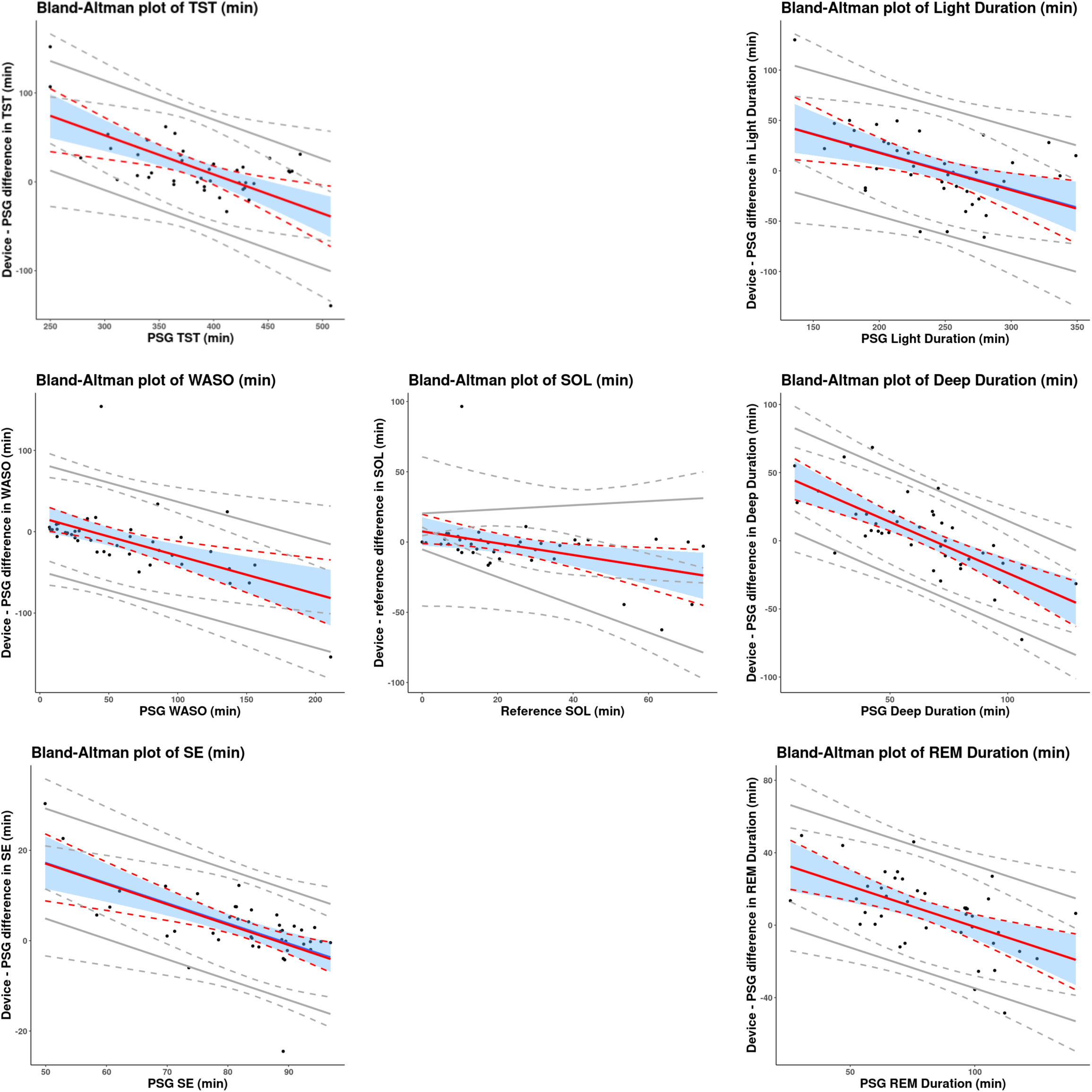
Bland-Altman plots of derived sleep measures. Note: Solid red lines indicate mean bias. Dotted red lines indicate 95% CI of mean bias. Solid gray lines indicate the 95% LOAs. Dotted gray lines indicate 95% CI of LOAs. Black dots are observations. Abbreviations: CI= confidence interval; LOA = limits of agreement; REM = rapid eye movement; SD = standard deviation; SE = sleep efficiency; SOL = sleep onset latency; TST = total sleep time; WASO = wake after sleep onset

**Table 3.**
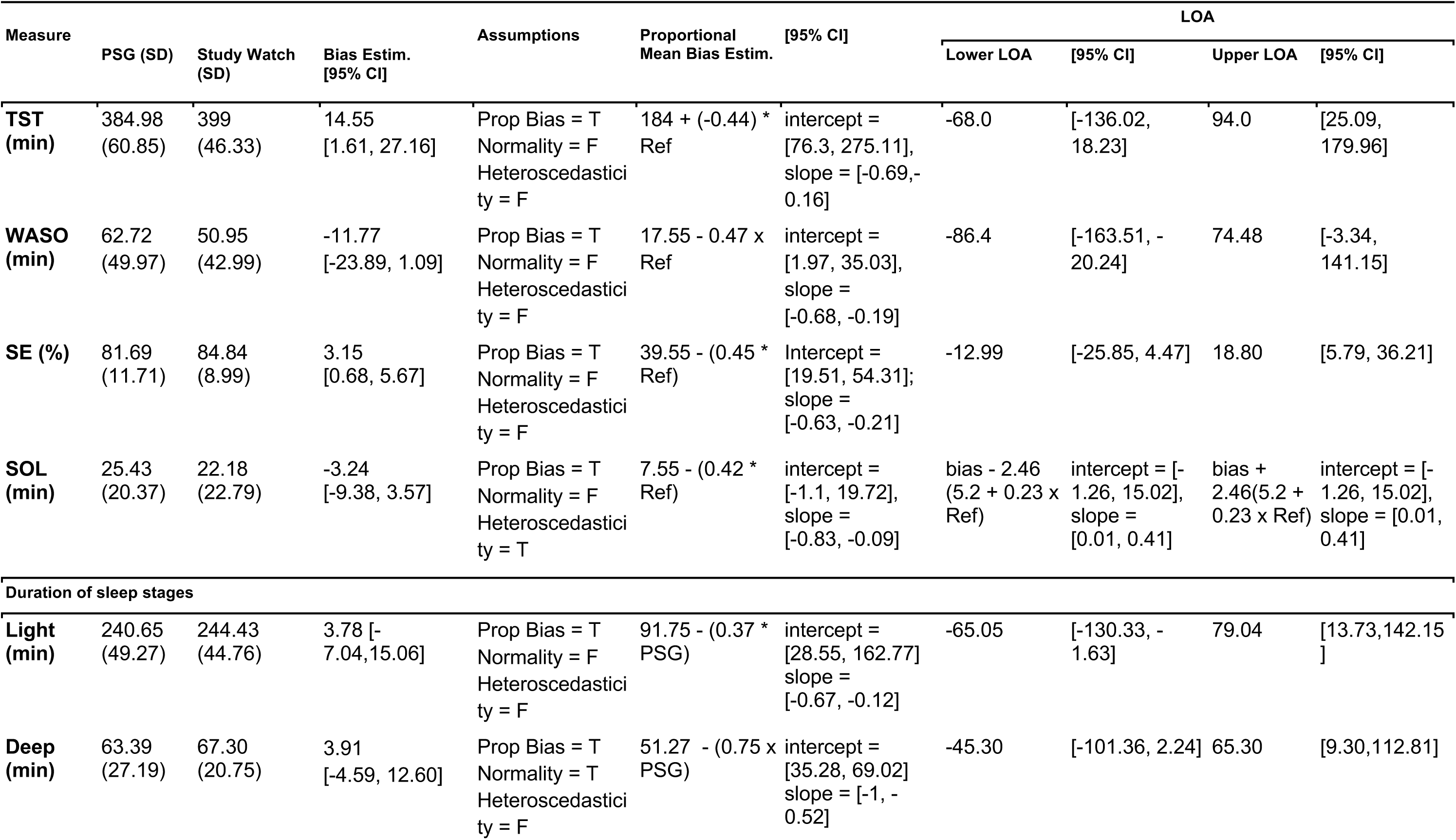
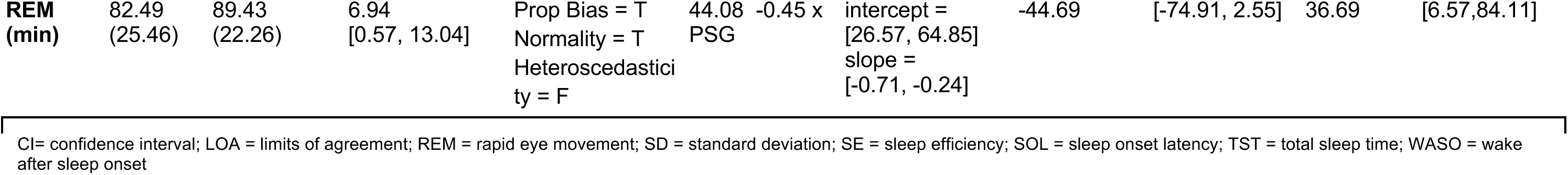
Summary of performance metrics for derived sleep measures.

The overall accuracy of the VNW algorithm in classifying sleep stages was 0.78 (95% CI: 0.51, 0.88), and the overall kappa was 0.64 (95% CI: 0.08, 0.82) (Table 4). There was variability in the performance across different sleep stages, with light sleep stage prediction having the lowest accuracy (Table 4), as there were instances of confusion between light sleep stage and all other stages (Table 5).

**Table 4.**
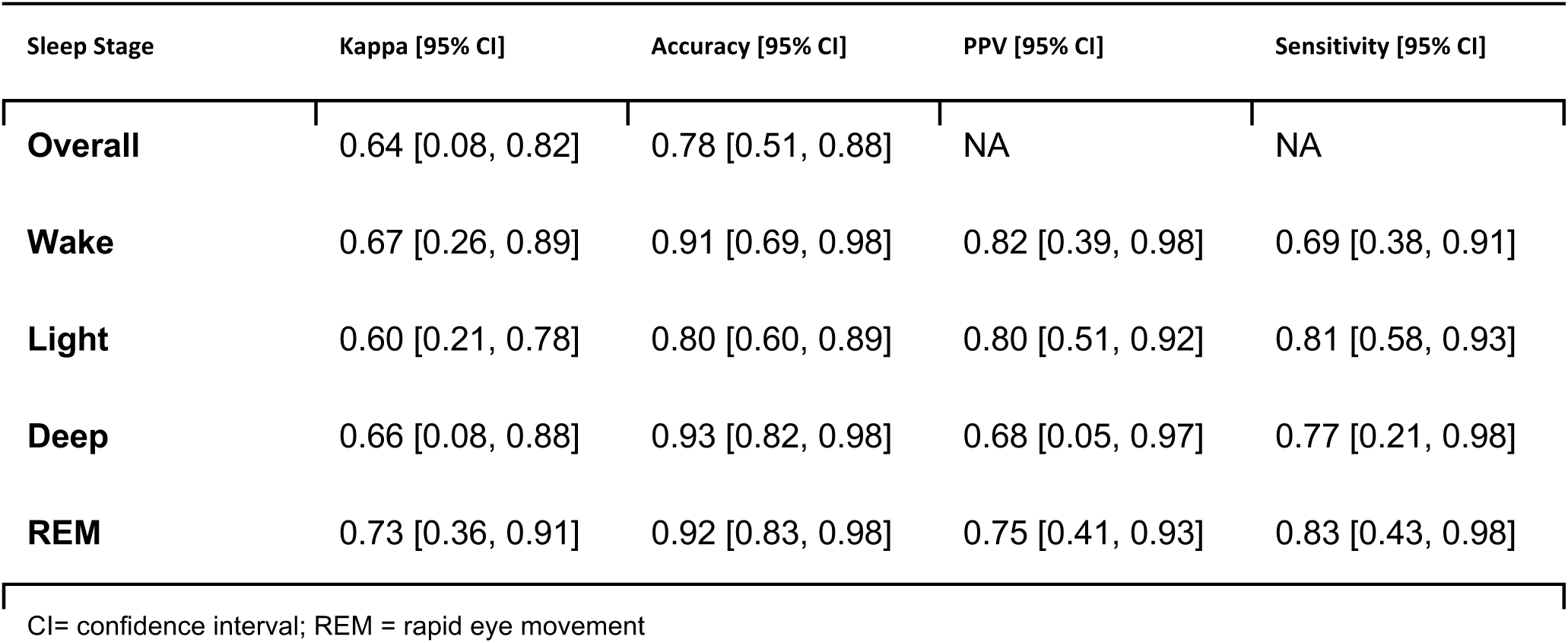
Summary of performance metrics for the classification of sleep stages.

**Table 5.**
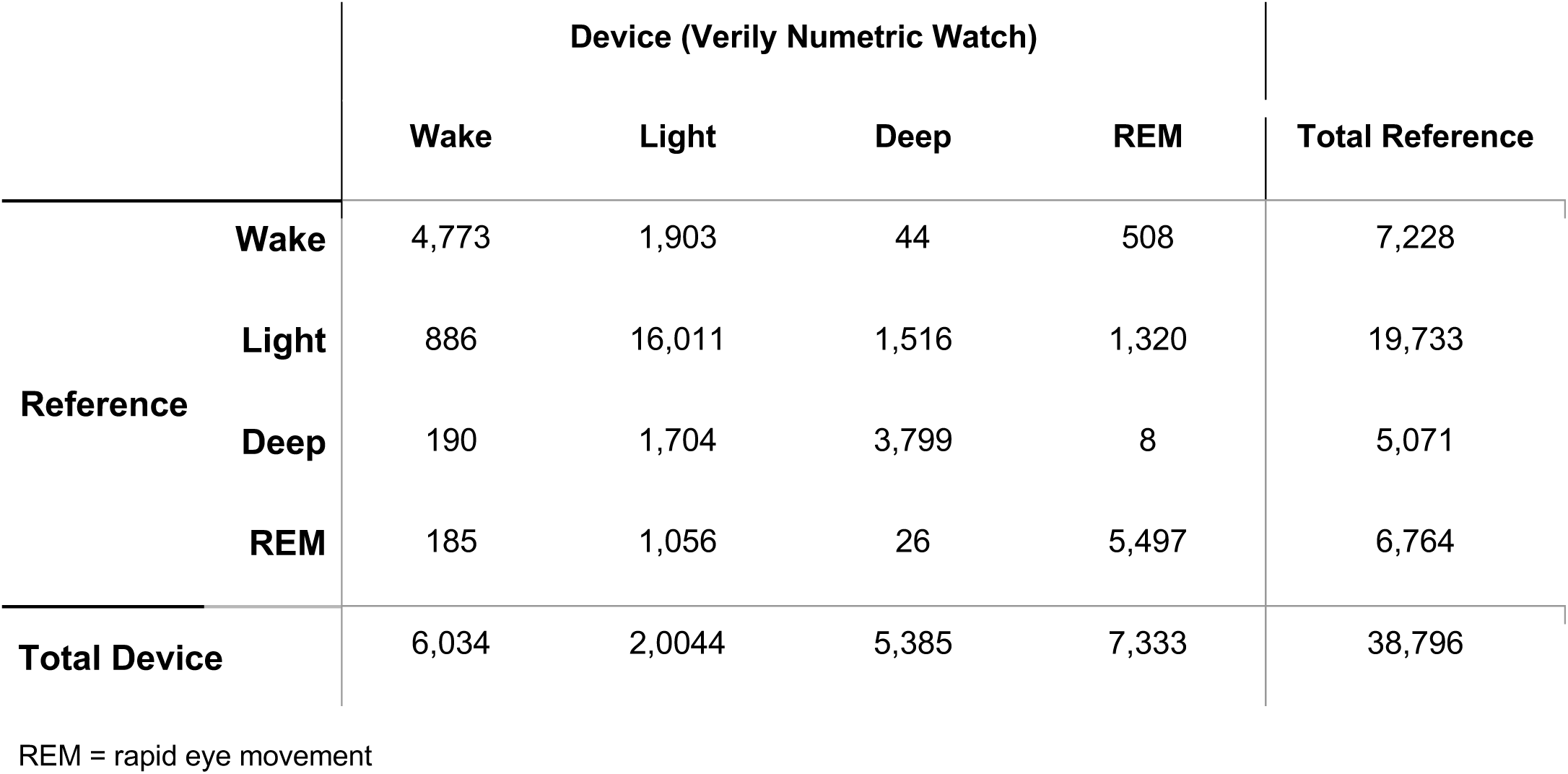
Confusion matrix for the classification of sleep stages.

|  |  | Device (Verily Numetric Watch) |  |  |  | Total Reference |
| --- | --- | --- | --- | --- | --- | --- |
|  |  | Wake | Light | Deep | REM |  |
| **Reference** | **Wake** | 4,773 | 1,903 | 44 | 508 | 7,228 |
|  | **Light** | 886 | 16,011 | 1,516 | 1,320 | 19,733 |
|  | **Deep** | 190 | 1,704 | 3,799 | 8 | 5,071 |
|  | **REM** | 185 | 1,056 | 26 | 5,497 | 6,764 |
| **Total Device** |  | 6,034 | 2,0044 | 5,385 | 7,333 | 38,796 |

REM = rapid eye movement

### Additional analytics and exploratory analyses

Subgroup analyses of the performance for the sleep vs wake classification, as well as the derived sleep measures, revealed results largely consistent with the overall group (Figure 2; Supplement Tables 2-12).

**Figure 2.**
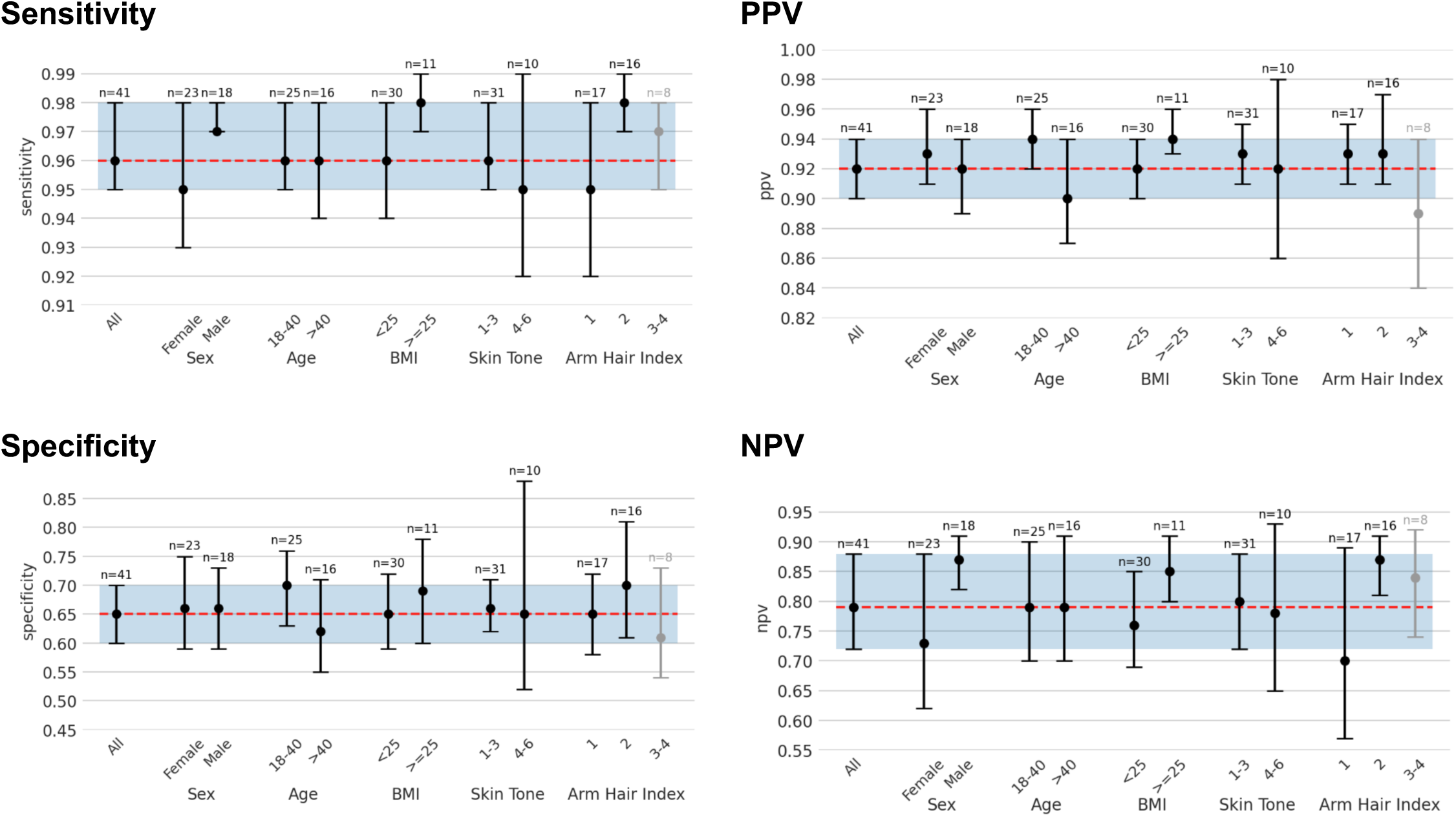
Performance of sleep vs wake classification across various subgroups based on sex, age, BMI, skin tone, and arm hair index. Notes: Bars show 95% CI ranges; dots, point estimates. For reference: red dotted line, overall point estimate; blue shade, overall 95% CI. For the skin tone classification, larger values indicate darker skin tone; for Arm Hair Index, larger values indicate coarser arm hair. Gray bars indicate subgroup with sample size < 10 participants. Abbreviations: BMI = body mass index; PPV = positive predictive value; NPV=negative predictive value

## Discussion

This evaluation of the performance of the VNW’s algorithm-derived sleep measures compared to PSG epoch-by-epoch in sleepers without elevated insomnia symptoms or OSA showed that sleep versus wake classification performance estimates were largely comparable or numerically higher than previously published results for other commercial wearable devices.^34,35^

Similar to other novel wearable devices, the ability to provide a 4-stage sleep classification was another strength of the VNW as some actigraphy-based devices only allow for 2-stage sleep classification, which precludes the detection of specific sleep stages that are indicated in various cognitive functioning and disease processes, such as depression and Parkinson’s Disease.^36–40^

Overall accuracy of the VNW for sleep stage classification was similar to other studies for the 4-stage model on a sample without OSA and heightened insomnia symptoms (see Schyvens et al.^22^ and Chinoy et al.^23^ for example). There were a few participants that had a low number of epochs in a particular sleep stage, including one participant with no deep sleep epochs. The sparsity of epochs in a particular sleep stage caused highly variable Kappa estimates, which likely led to a wide 95% CI. We found VNW to show significant heteroskedasticity for SOL and significant proportional bias for TST, WASO, SE, SOL, and light, deep, and REM sleep duration, overestimating and underestimating shorter and longer values, respectively, both of which are common in other wearable devices.^41^ Based on a mean of the bias estimates (Table 3), the 95% CI for TST, WASO, and SOL biases fell within the range of allowable differences for actigraphy in clinical populations recommended by the AASM clinical practice guideline. However, when using the proportional mean bias estimate, which accounts for variation in bias over the range of measurement, the 95% CI exceeds the allowable difference at lower and higher ends of the distributions (Figure 1, Table 3). These allowable differences are also based on adults with specific sleep disorders and the current sample did not include patients with these characteristics.

Investigating the generalizability of the performance of the VNW among different demographic subgroups was an aspect of special interest in our study. VNW performed consistently across subgroups for the classification of sleep vs wake and sleep stages, albeit with larger variability in estimates for those with darker skin tone. In addition, for the calculated sleep measures, all mean bias 95% CIs overlapped across demographic subgroups (i.e., sex, age, BMI, skin tone, arm hair density), indicating that there were likely no statistically significant differences between groups; but as stated previously, limited sample sizes may preclude the ability to detect differences between groups. Lastly, a slight difference in performance was observed between age groups (younger vs older) and between sexes.

### Limitations and future perspectives

Strengths of this study include thoroughness of statistical methods,^19^ the compliance with state-of-the art recommendation for performance evaluation studies,^7^ and the diverse sample of participants, with a range of ages, BMI, skin tones, and arm hair density, which are known factors to influence PPG signal quality. Nonetheless, there are important limitations to consider in our study.

First, we focused this investigation on data collected from a sample of adult sleepers without elevated insomnia symptoms or OSA. In order to fully characterize performance in clinically relevant scenarios, future studies should evaluate the performance of this device in populations with disturbances in sleep, such as people with sleep disorders. Similarly, although our study group had about 30% of participants with overweight/obese BMI, it is unknown whether performance would generalize to morbidly obese populations, whose watch fit and tissue characteristics may vary and who often have sleep apnea as a comorbidity.

Second, as a typical procedure for the performance evaluation of sleep measures with in-lab PSG as the reference standard, epoch by epoch accuracy evaluation was conducted between lights-off and lights-on. This may limit the interpretation or generalizability of SOL results, as there is no way to easily passively identify the moment a participant begins to attempt to initiate sleep in real-world settings. In addition, as is common practice, participants started wearing the VNW before PSG was on, the start of the PSG recording was labeled as “Time 0”, and time was rounded (e.g., 22:32:00).^42^ Yet, the mean overall difference between the onset times of the devices was below the range shown to introduce significant bias in study outcomes (see Supplementary Materials). While there is always some degree of error introduced by alignment methods, we cannot rule out that this may have introduced some error that would have resulted in a small degree of underestimating performance of staging.

Third, our subgroup analyses were underpowered, limiting our ability to extract robust conclusions from them. Future studies should aim to recruit larger diverse cohorts, particularly sampling for specific key features that may affect device performance, including but not limited to skin tone, arm hair density, and clinical status.

## Conclusion

Results demonstrate the potential of the VNW to effectively measure 12 standard sleep metrics, as compared to gold-standard PSG-based labels, in a demographically diverse sample of adults. Results from the epoch by epoch sleep versus wake classification, sleep stage classification, and from the derived overnight sleep measures showed comparable performance across demographic subgroups. The results support the application of this device to monitor and understand sleep behaviors in sleepers without elevated insomnia symptoms or OSA in free-living settings for long durations, when PSG collection is not an optimal method.

## Data Availability

Data from this study are not available due to the nature of this program. Participants did not consent for their data to be shared publicly.

## Supplement

### Detailed description of enrollment screening

Questionnaires completed by participants or study personnel and used for screening were:

- Obstructive Sleep Apnea 50 (OSA50): a brief 4-item (obesity, snoring, apneas, and age) diagnostic screening questionnaire for OSA in primary care that has been shown to identify patients with moderate to severe OSA with 84% accuracy ^24^. This questionnaire has a cut-off score of ≥ 5, with typical sleepers having an OSA50 score of < 5 (sensitivity 100%, specificity 29%).
- AHI: calculated from PSG signals, it is the total number of apnea or hypopnea events in a night divided by the hours of sleep. The AHI assists in the diagnosis of OSA, classifying participants as having Normal sleep (< 5 events per hour), Mild (5-14 events per hour), Moderate (15-29 events per hour), and Severe (30 or more events per hour).
- Insomnia Severity Index (ISI): a 7-item questionnaire designed to screen for insomnia ^25^. A 5-point Likert scale ranging from 0 = no problem to 4 = very severe problem is used for each question to calculate a total score ranging from 0 to 28. Total scores are categorized as absence of insomnia (0–7); sub-threshold insomnia (8–14); moderate insomnia (15–21); and severe insomnia (22–28). An ISI < 8 is used to identify no clinically significant insomnia symptoms.
- ESS: an 8-item self-reported questionnaire used to assess daytime sleepiness ^30^. A 4-point Likert scale ranging from 0 = would never doze to 3 = high chance of dozing is used for each question to calculate a total score ranging from 0 to 24. Total scores are categorized as normal (0-10) and then increasing degree of daytime sleepiness (11-24). This measure has good internal consistency^43^.
- Fitzpatrick Skin Scale (completed by study personnel) ^31^: a 10-item questionnaire that assesses Genetic (physical traits), Sensitivity (reaction to sun exposure), and Intentional Exposure (tanning habits) to categorize participants on a scale from 1-6 as research has shown that skin tone can influence the accuracy of PPG signals. Categories include, Type I (scores 0–6) always burns, never tans (palest; freckles), Type II (scores 7–13) usually burns, tans minimally (light colored but darker than fair), Type III (scores 14–20) sometimes mild burn, tans uniformly (golden honey or olive), Type IV (scores 21–27) burns minimally, always tans well (moderate brown), Type V (scores 28–34) very rarely burns, tans very easily (dark brown), Type VI (scores 35–36) never burns (deeply pigmented dark brown to darkest brown). These can be categorized into light (Type I and II), medium (Type III and IV), and dark (Type V and VI)^44^.
- Arm Hair Index (scored by study personnel): assesses the density of participants’ arm hair on a scale of one to four, including 1 (Little to no visible hair), 2 (Visible fine arm hair), 3 (Coarse arm hair), and 4 (Very coarse arm hair).

### Device synchronization during overnight study visit

Time synchronization between the VNW and PSG devices was performed using the following steps. ^42^ First, the VNW was placed on the non-dominant wrist and the time (HH:MM) was recorded in the Device Accountability Log and electronic data capture (EDC) system.

Next, PSG hook up and calibration were performed; PSG was then turned on at the top of the minute on the VNW device (i.e., when the minute on the watch face changed) and the time (HH:MM) was logged in the EDC system.

Participants started wearing VNW before PSG was on and the overall difference between the onset times (mean = 3.51 seconds, SD = 4.06 seconds) was well below the range that has been shown to introduce significant bias in study outcomes. As is common practice, the start of the PSG recording was labeled as “Time 0” and time was rounded (e.g., 22:32:00).^42^

### Success Criteria for Sleep Versus Wake Classification as Product Requirement Specification

We pre-specified thresholds as “success criteria” for sleep versus wake classification with success being defined as sensitivity ≥ 0.90 and specificity ≥ 0.50 (the lower 95% CI bound over these values) as internal criteria for a successful sleep versus wake classification to proceed with subsequent development of the algorithm.

---

**Supplement Table 1.**
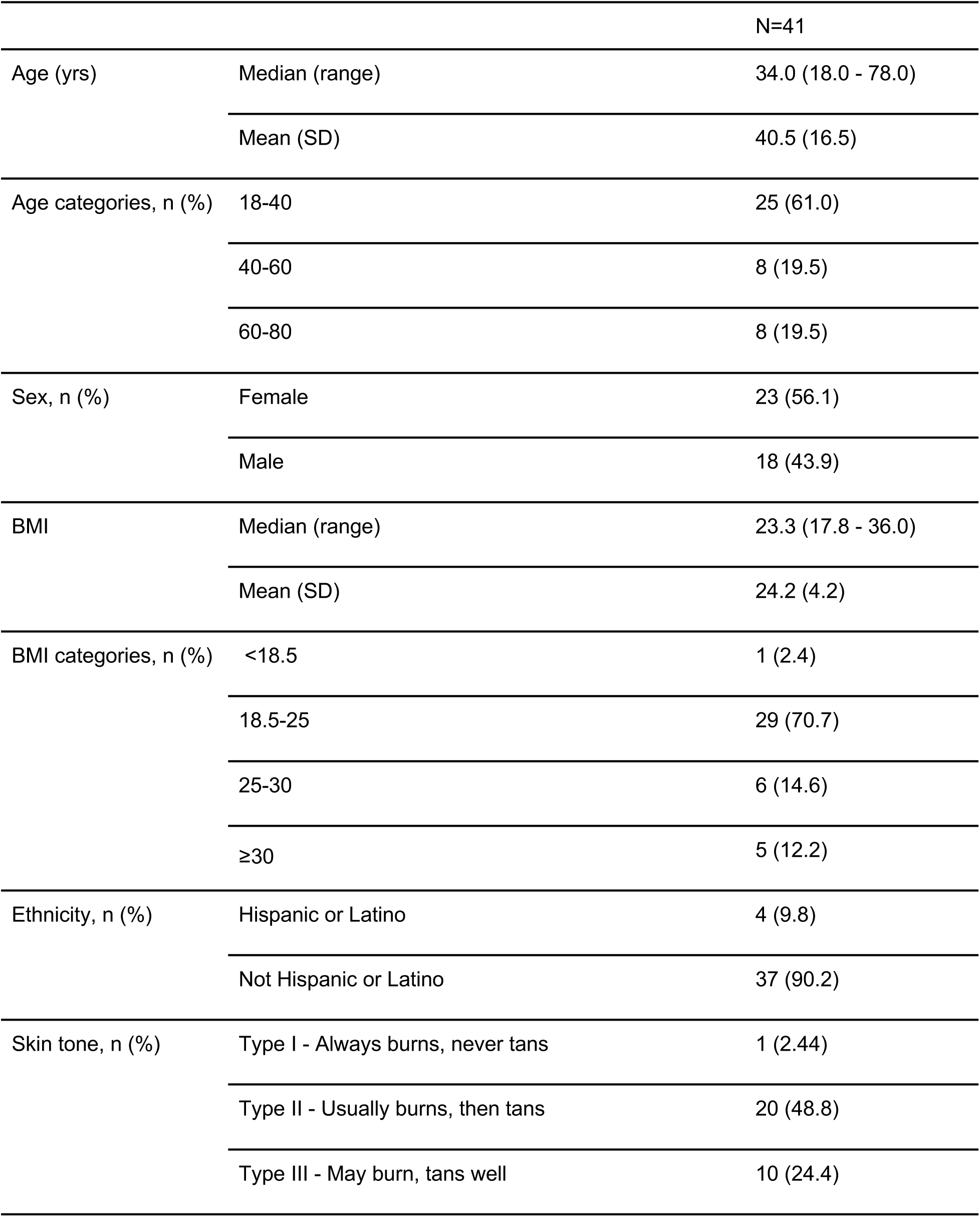
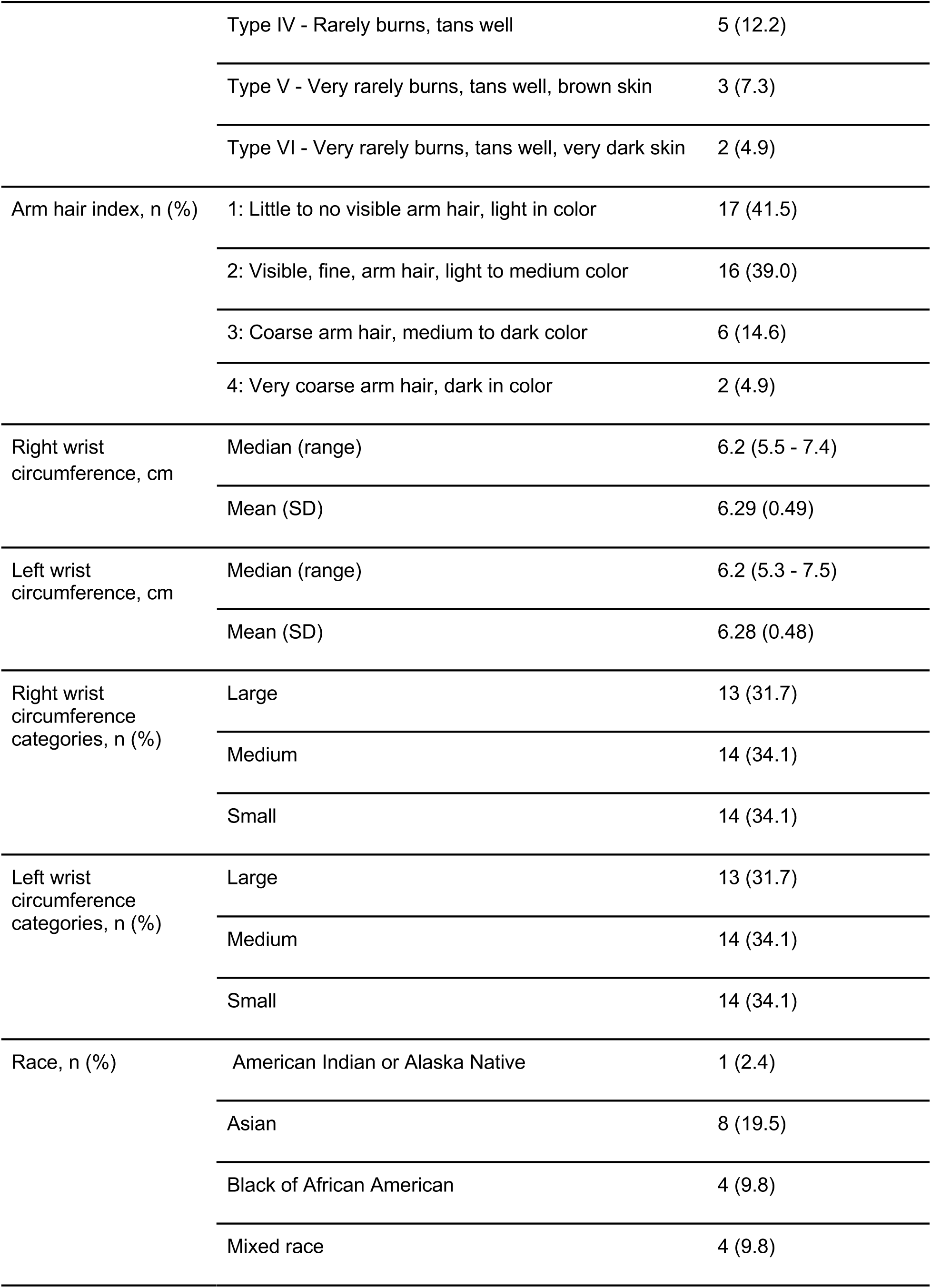
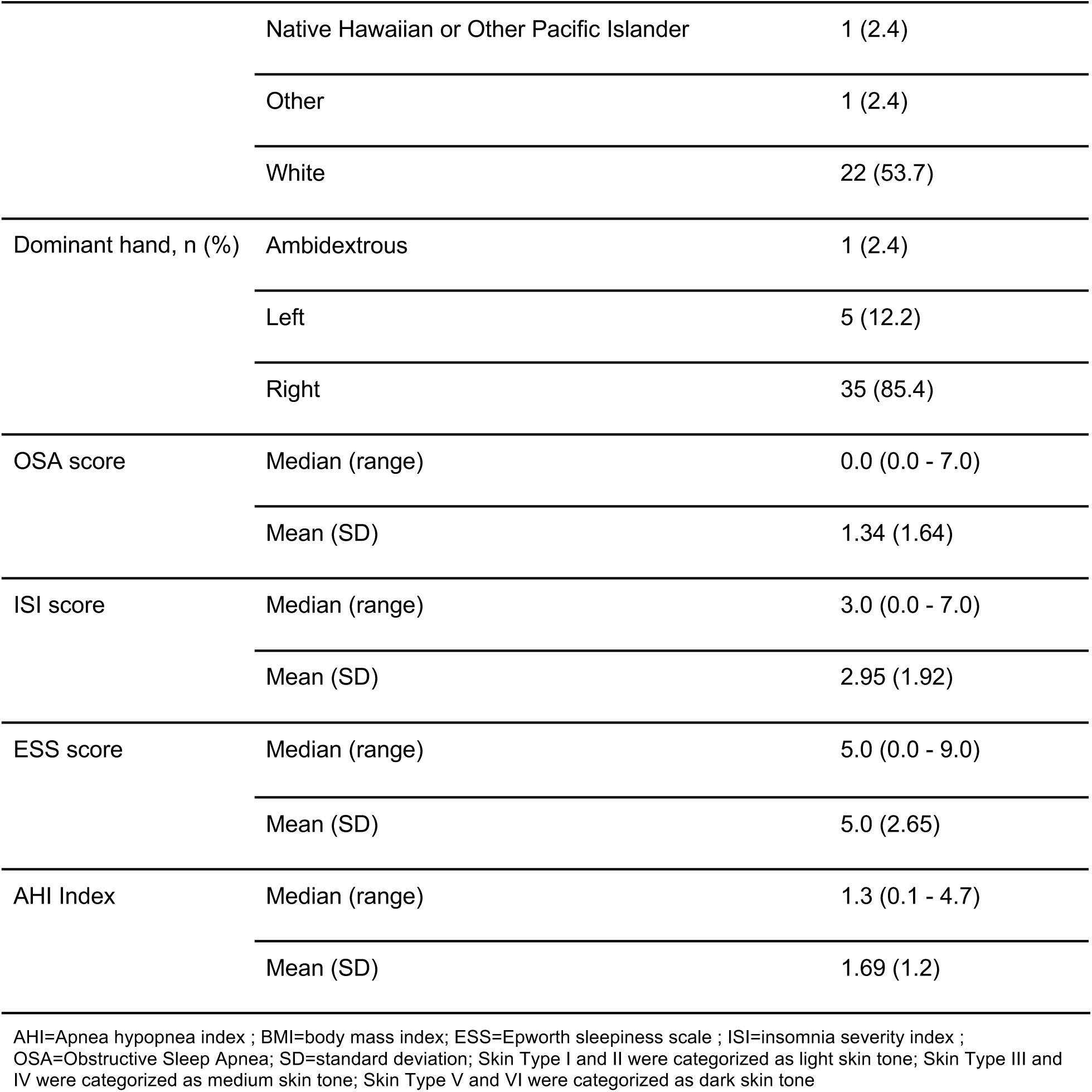
Full summary of participant characteristics.

**Supplement Table 2.**
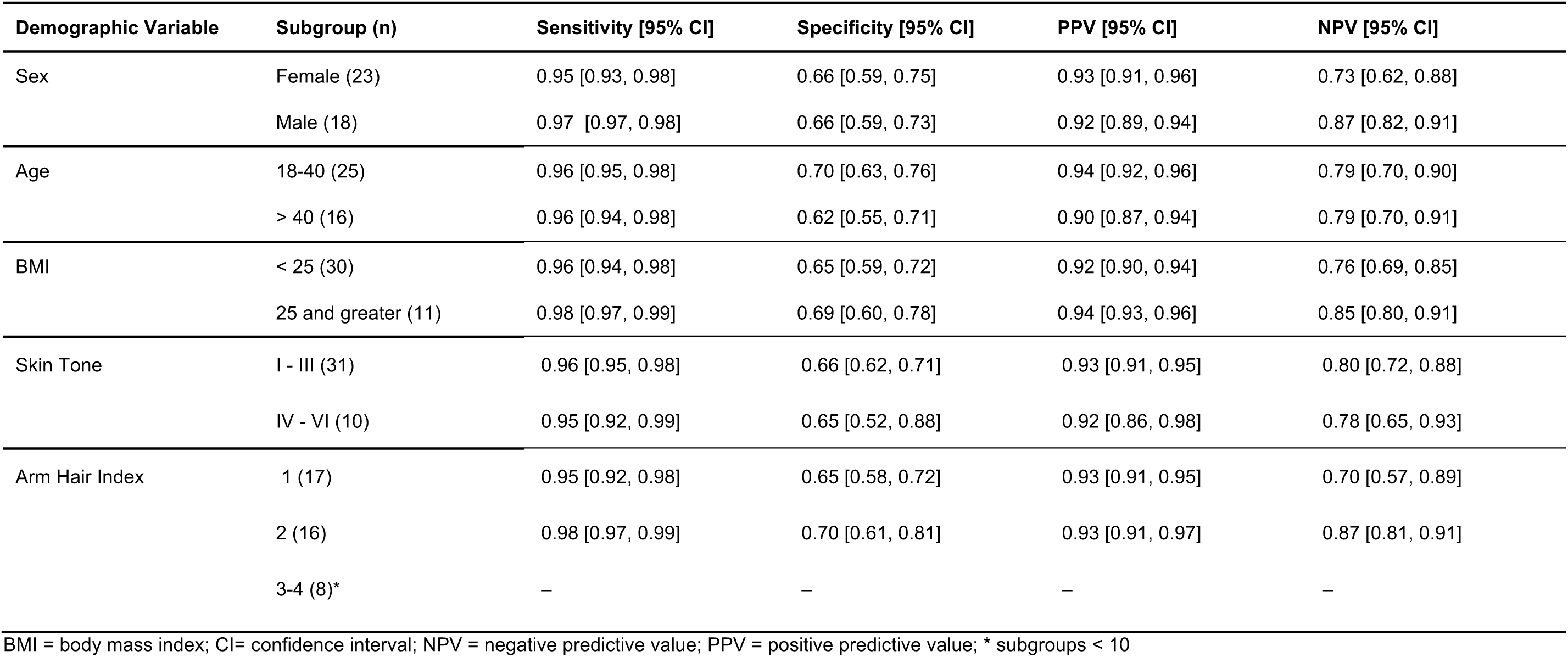
Summary of performance metrics for ‘sleep vs wake’ classification, according to participant subgroups.

**Supplement Table 3.**
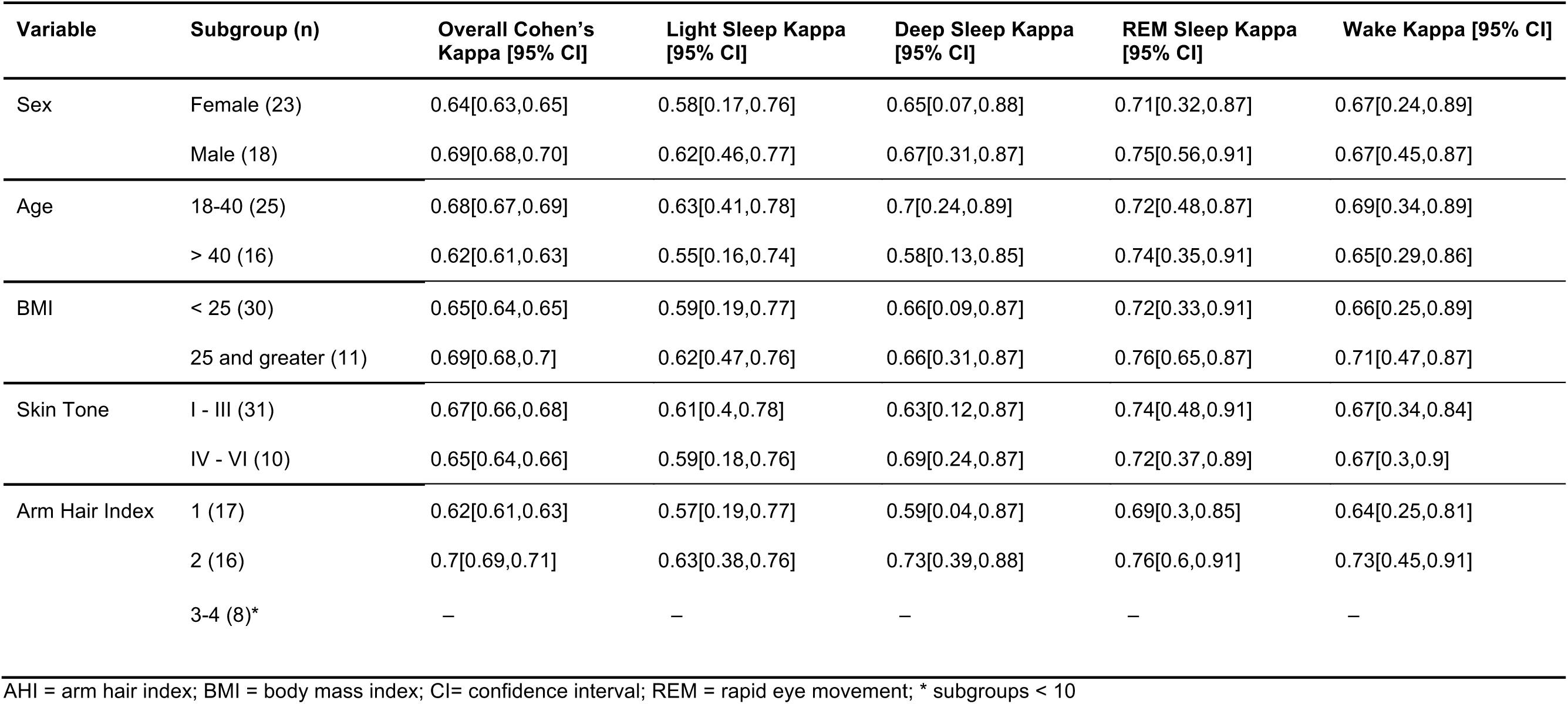
Summary of performance metrics for ‘sleep stage’’ classification, according to participant subgroups.

**Supplement Table 4.**
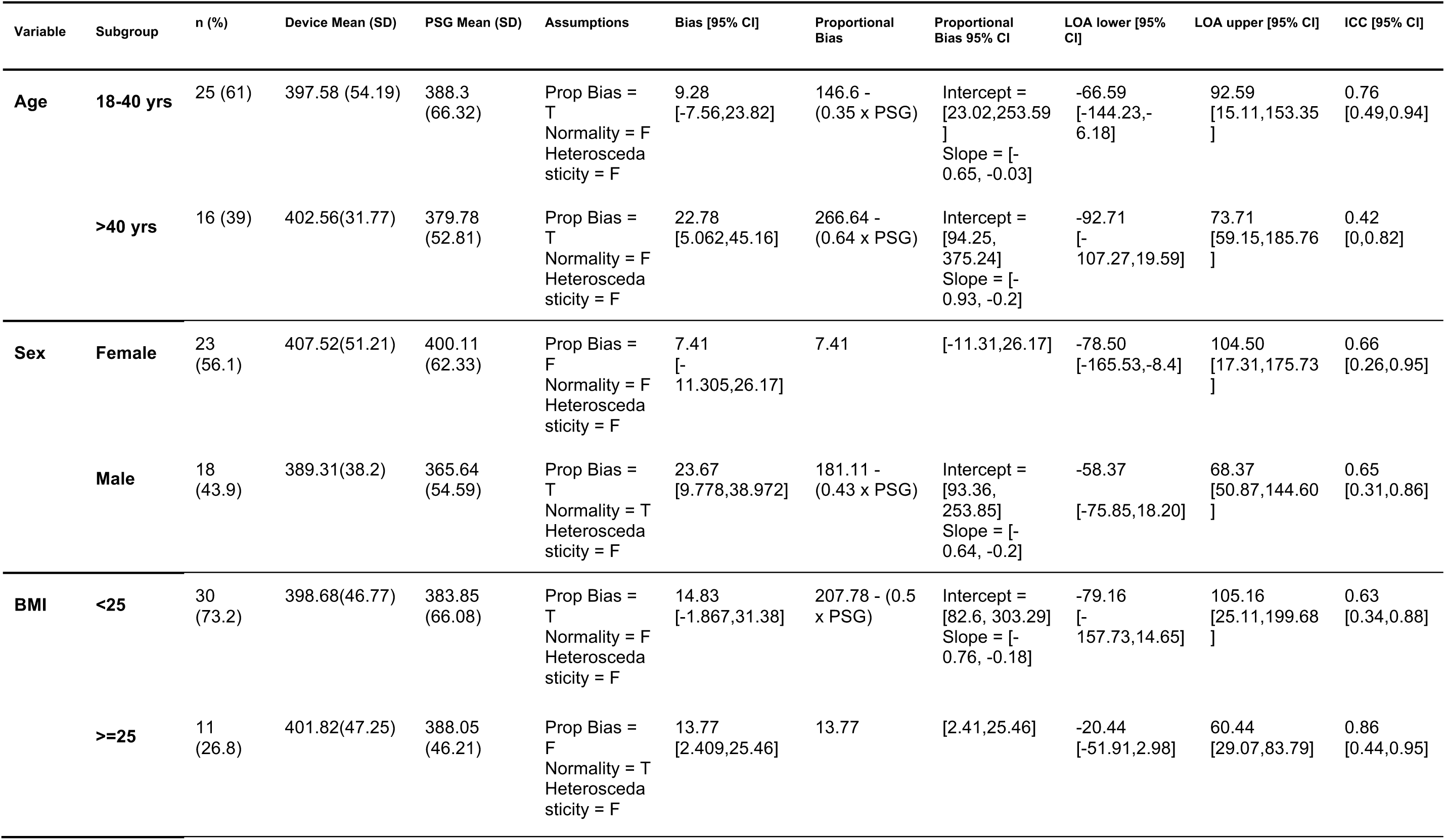
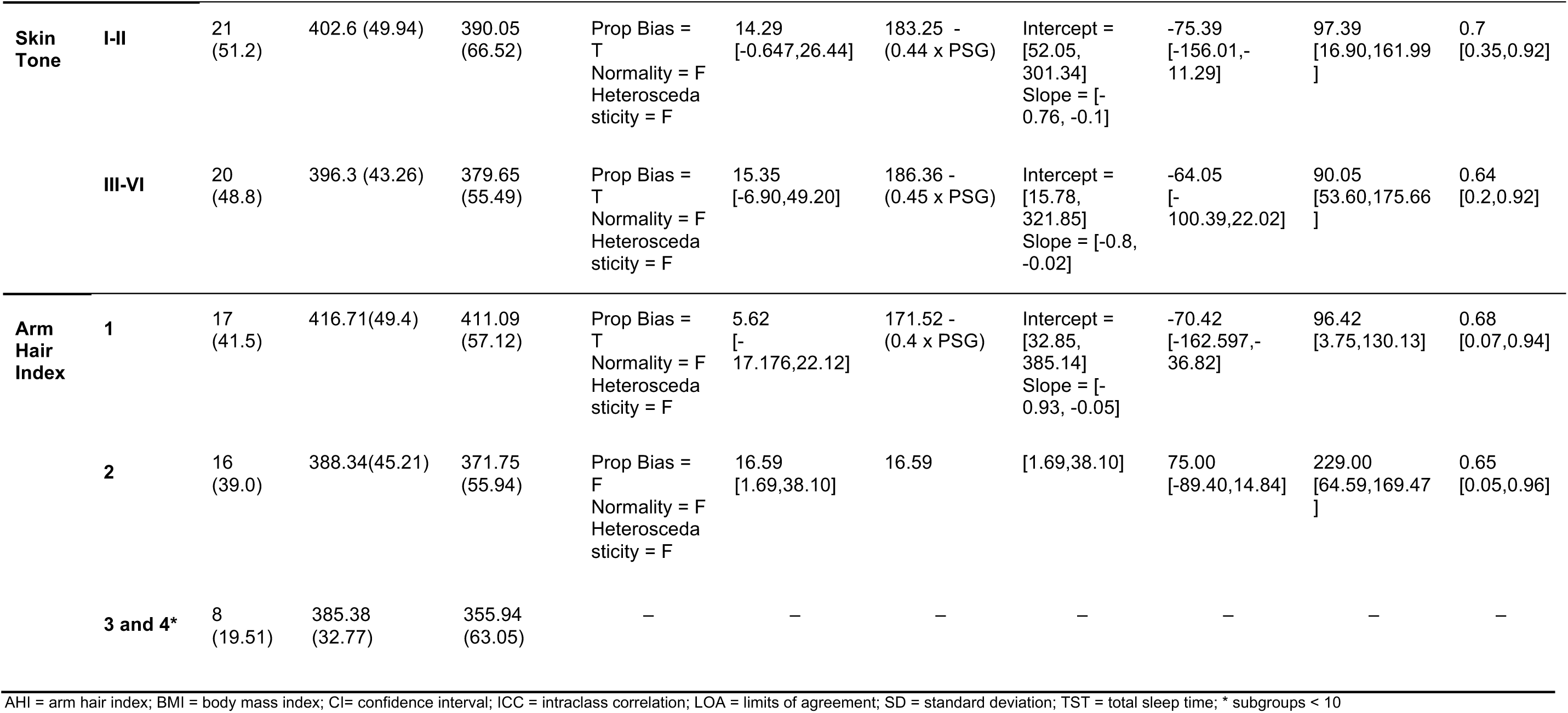
Summary of performance metrics for TST according to subgroups.

**Supplement Table 5.**
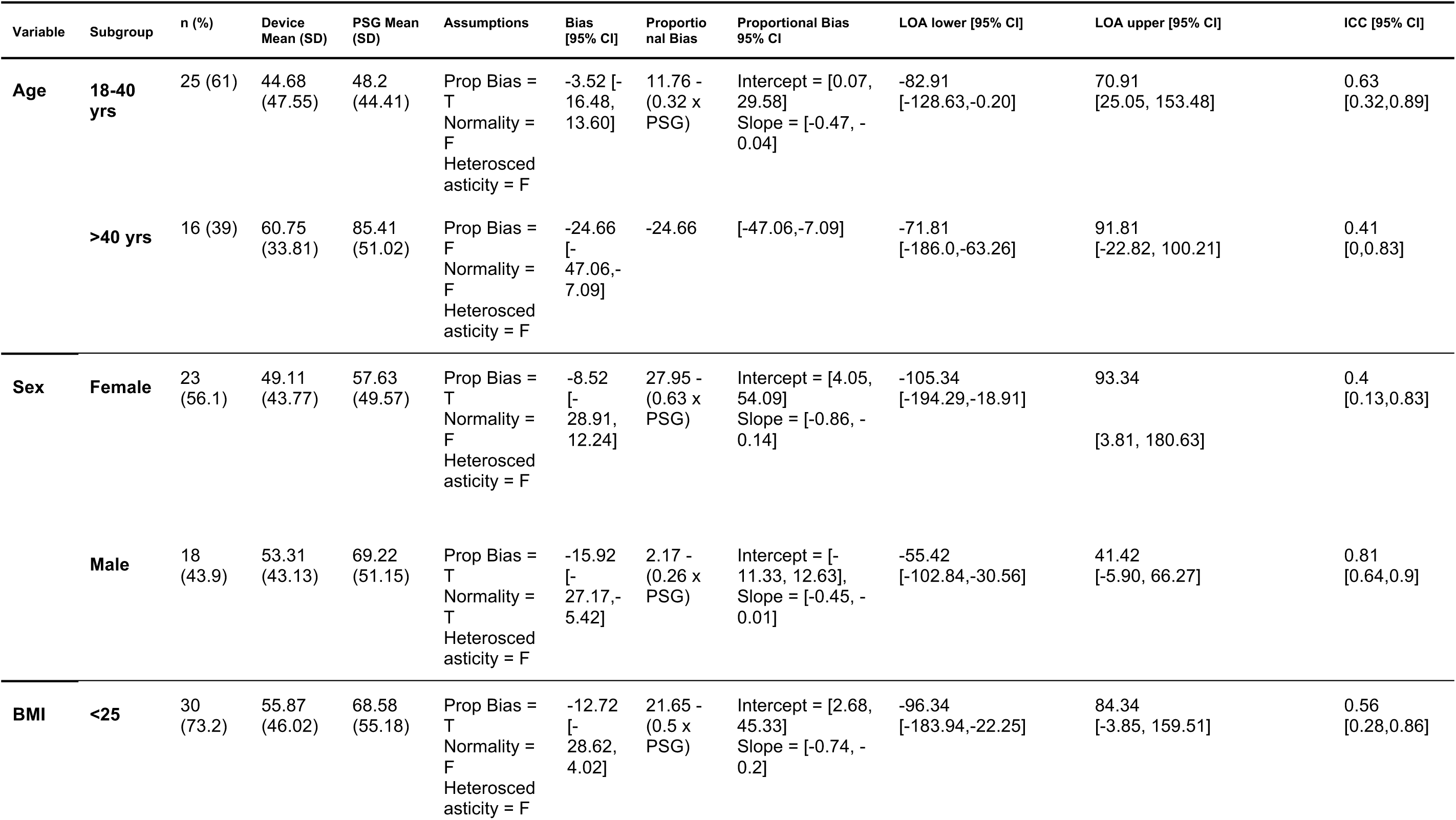
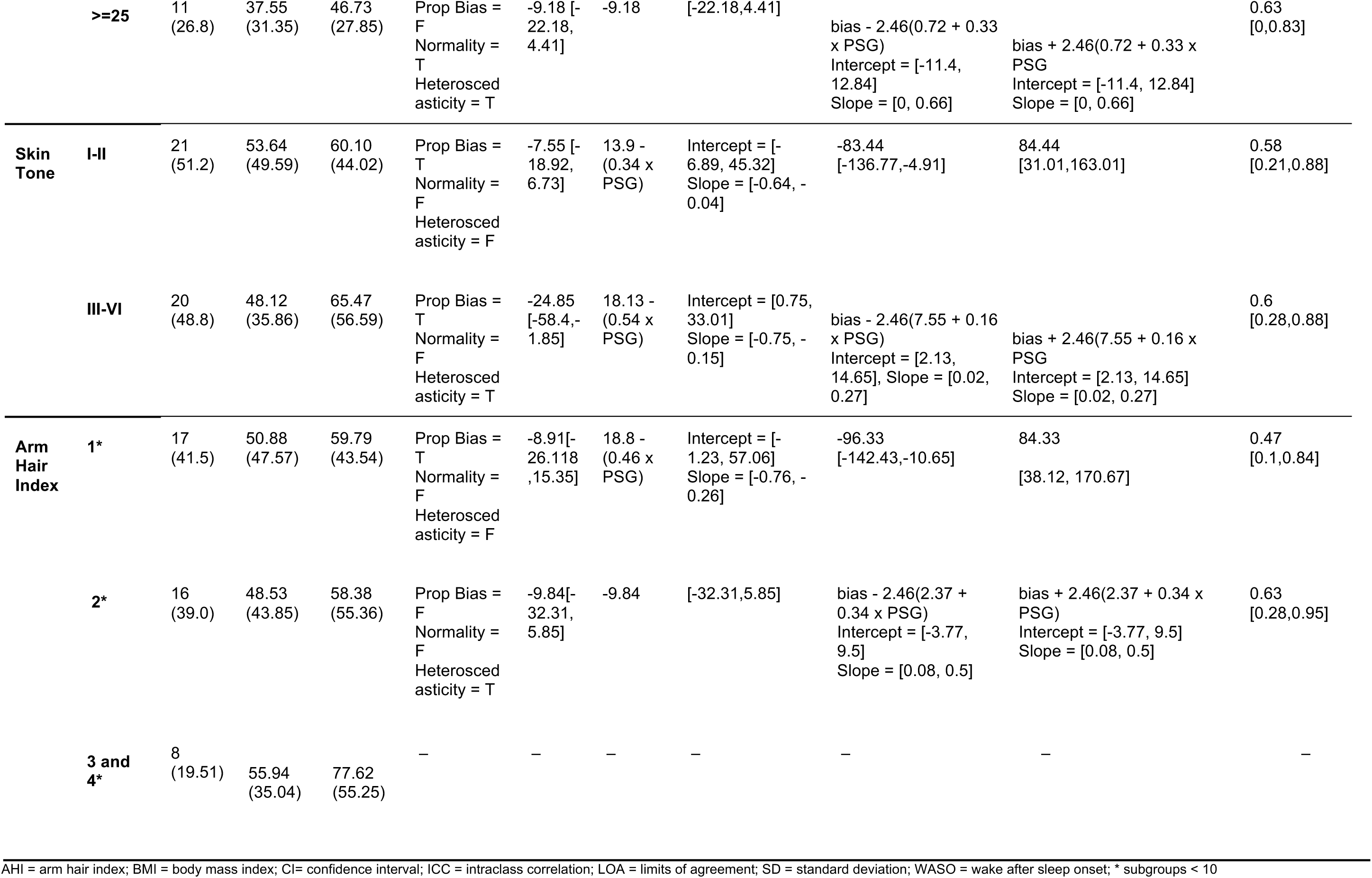
Summary of performance metrics for WASO according to subgroups.

**Supplement Table 6.**
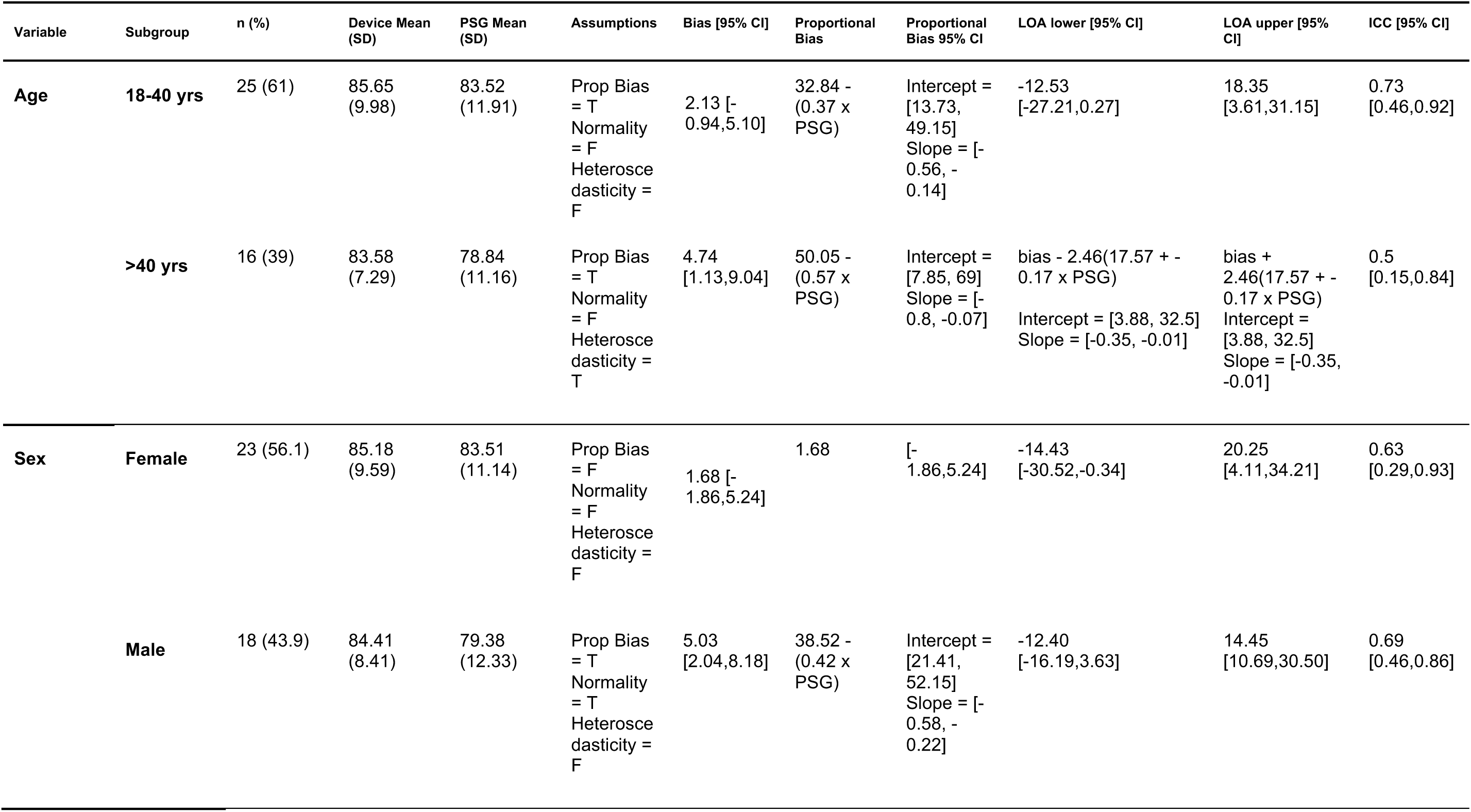
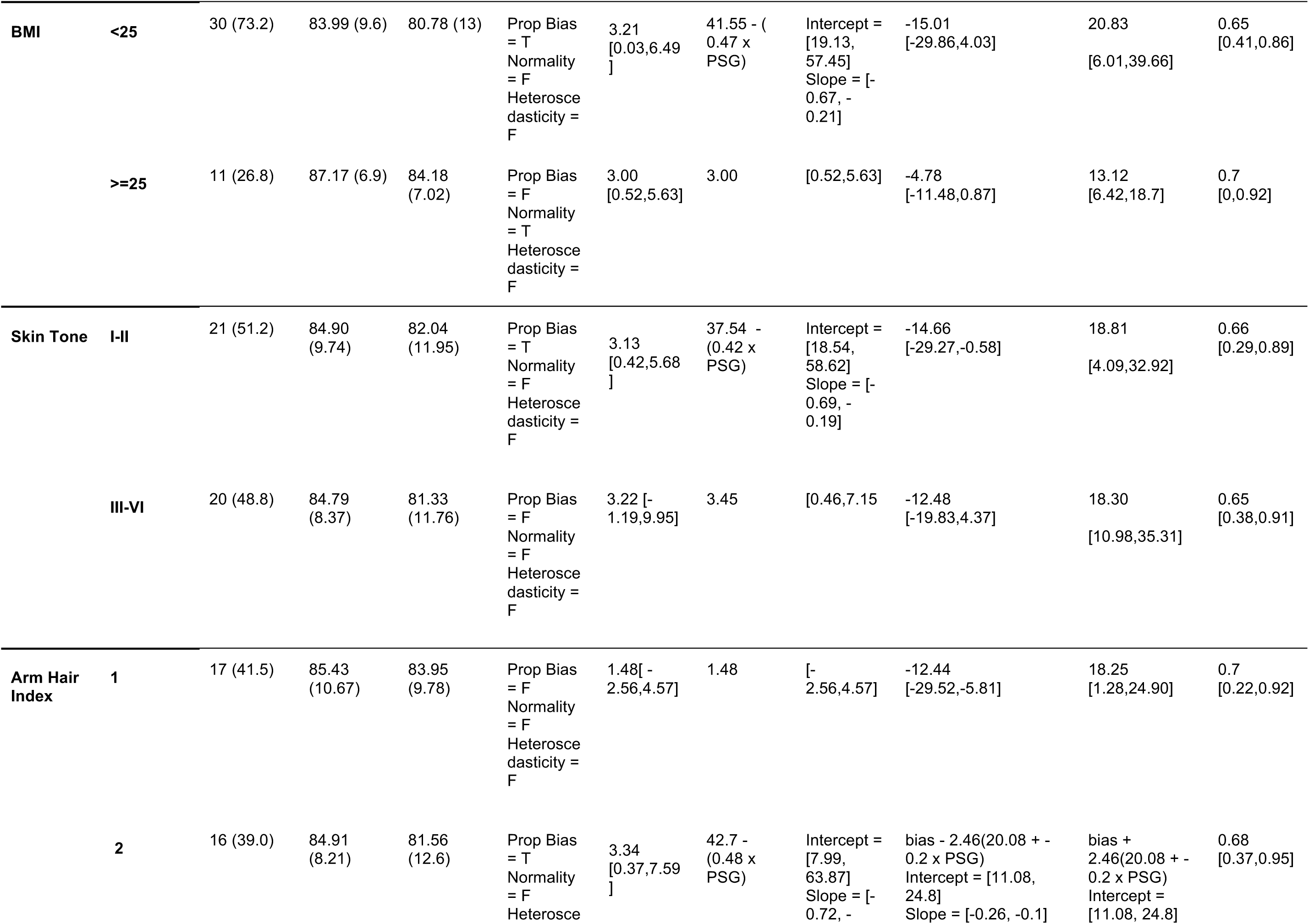
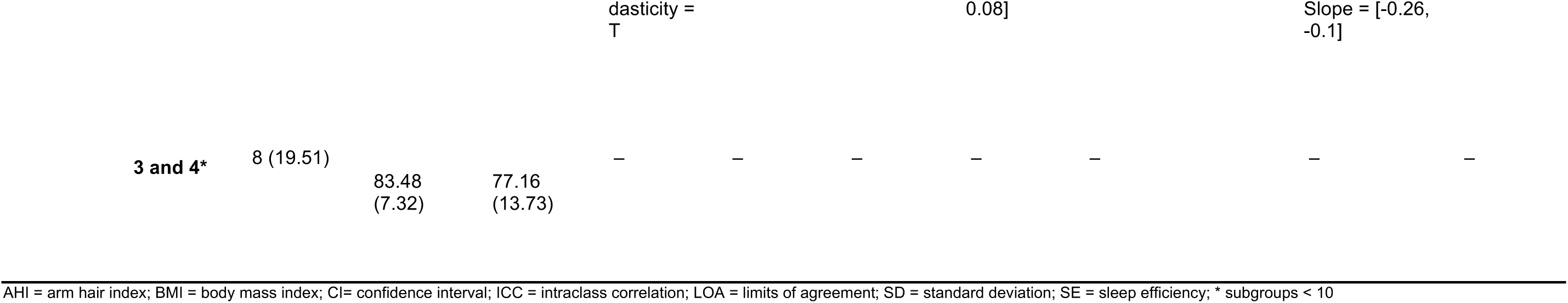
Summary of performance metrics for SE according to subgroups.

**Supplement Table 7.**
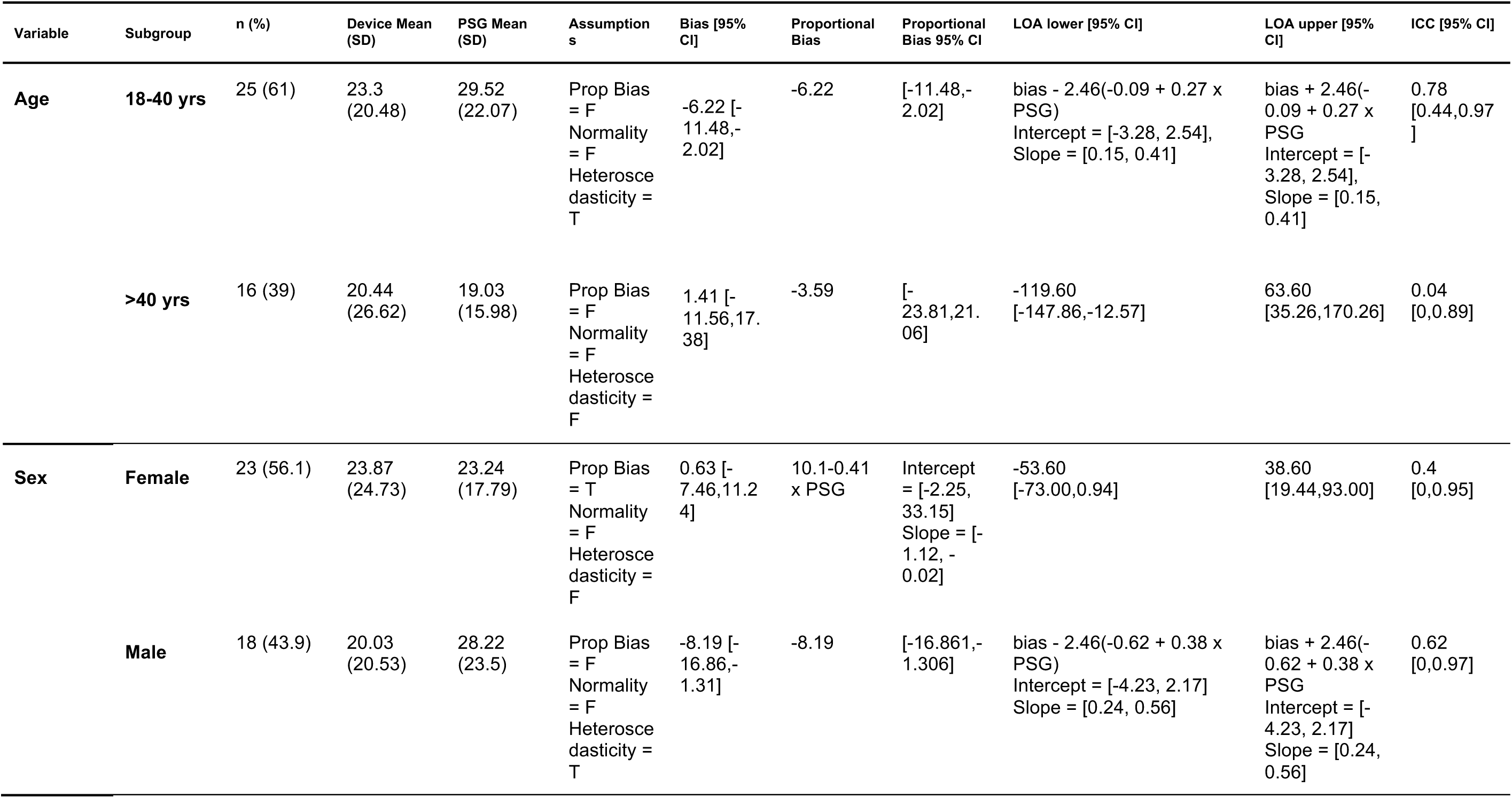
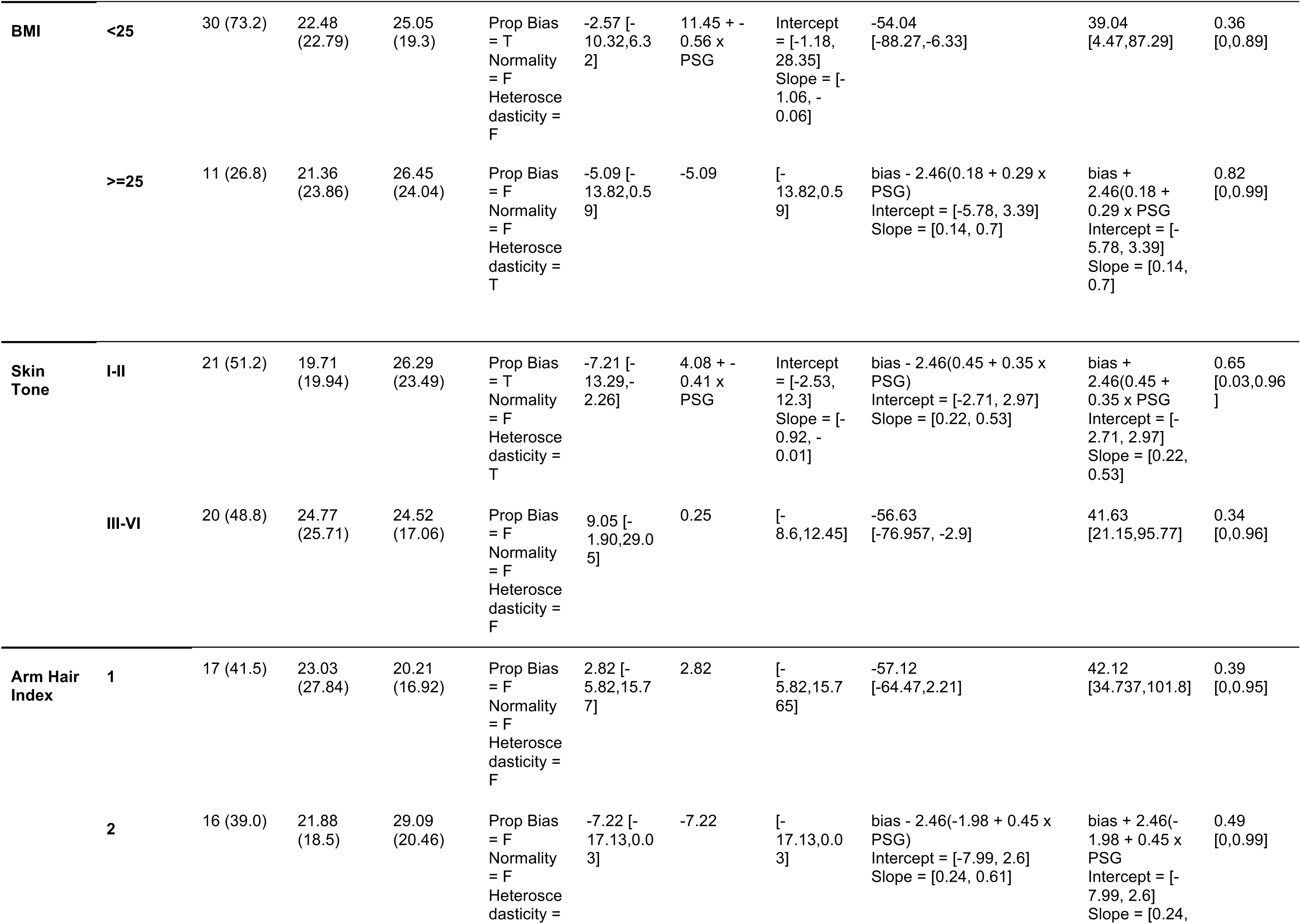
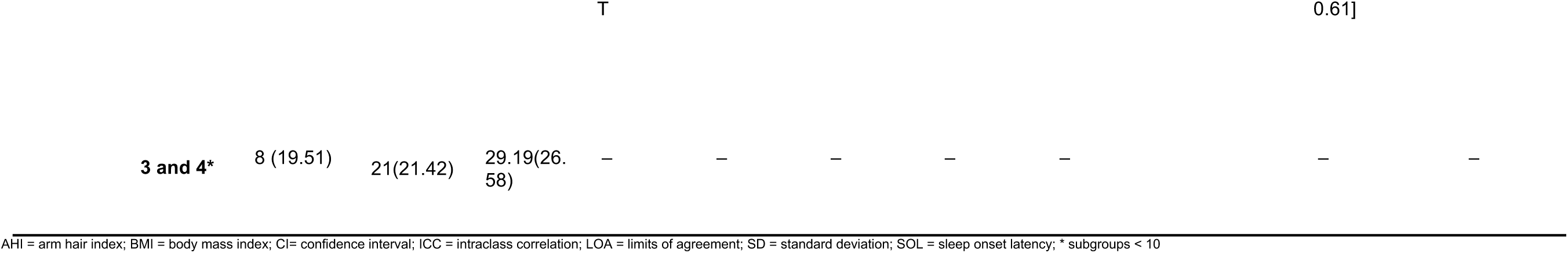
Summary of performance metrics for SOL according to subgroups.

**Supplement Table 8.**
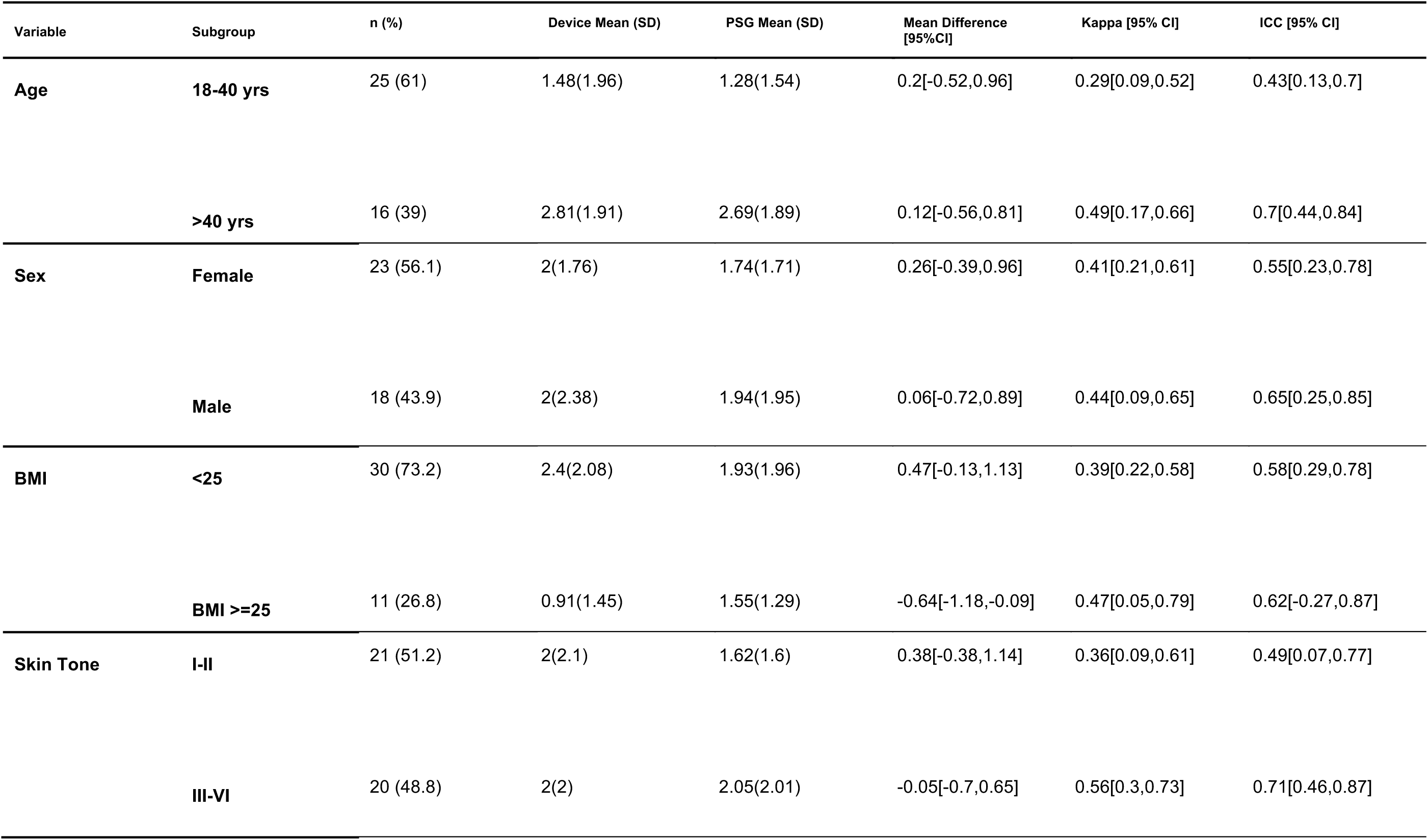
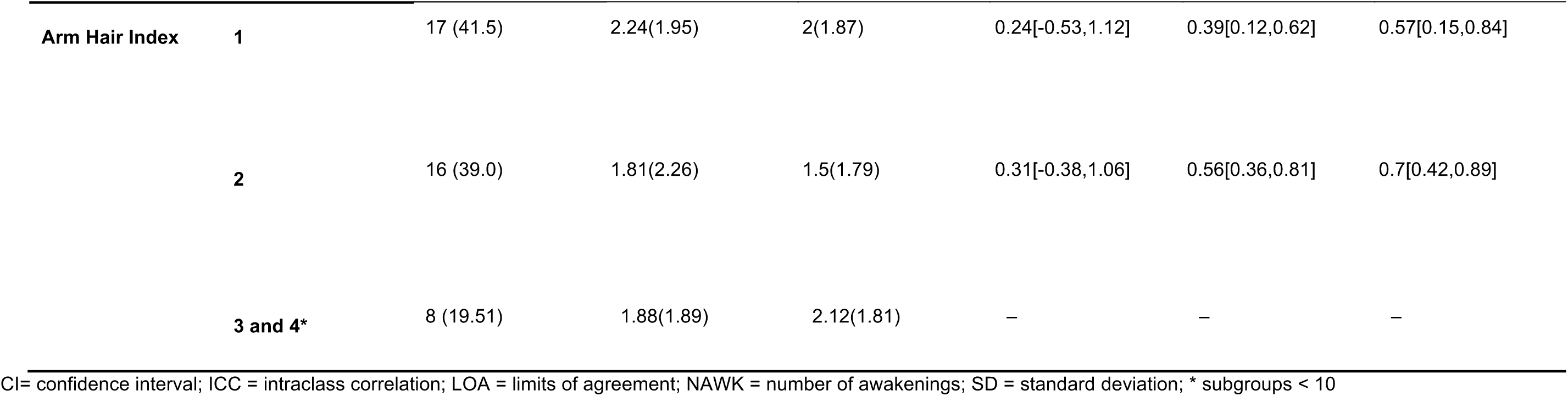
Summary of performance metrics for NAWK according to subgroups.

**Supplement Table 9.**
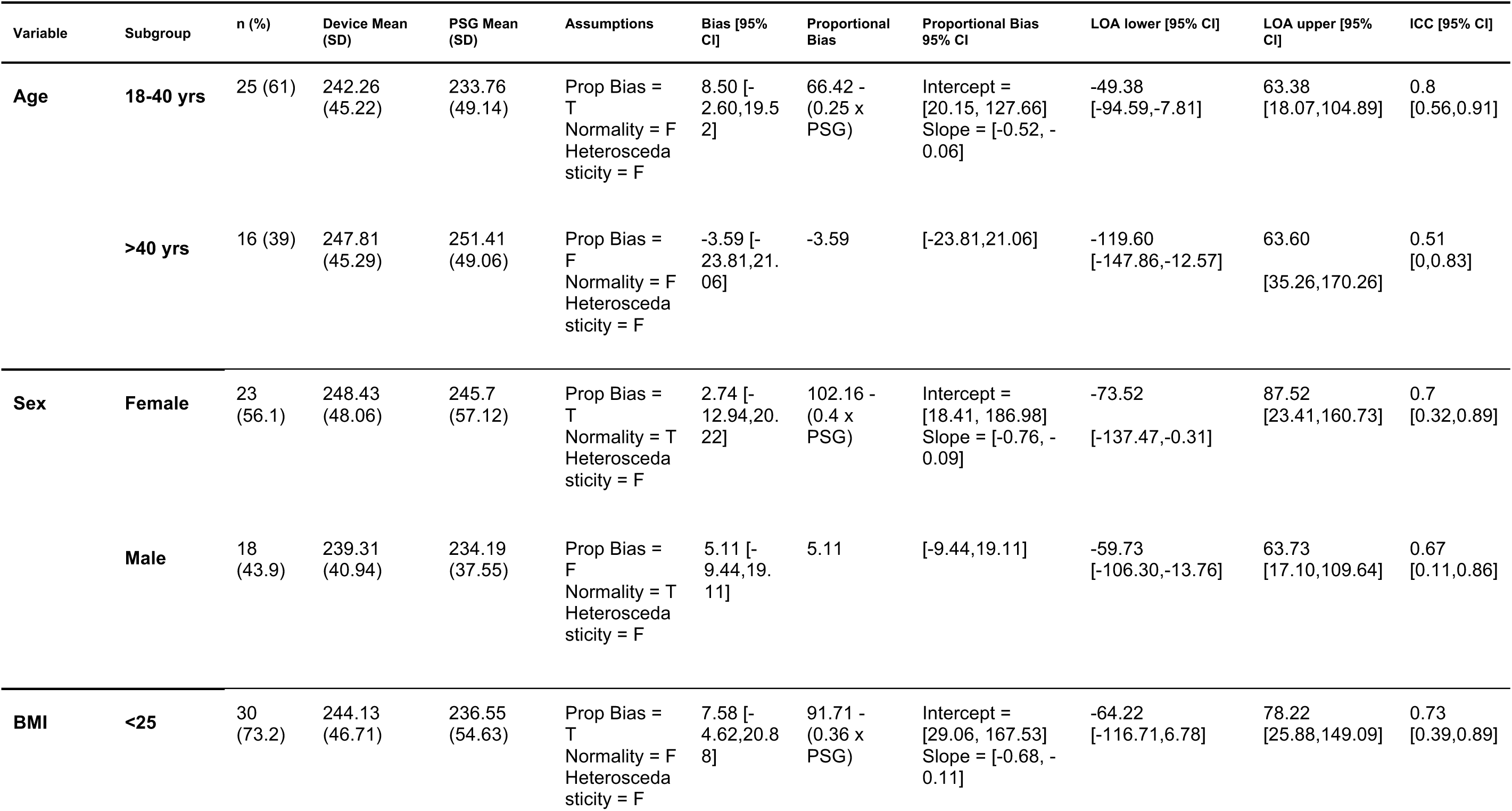
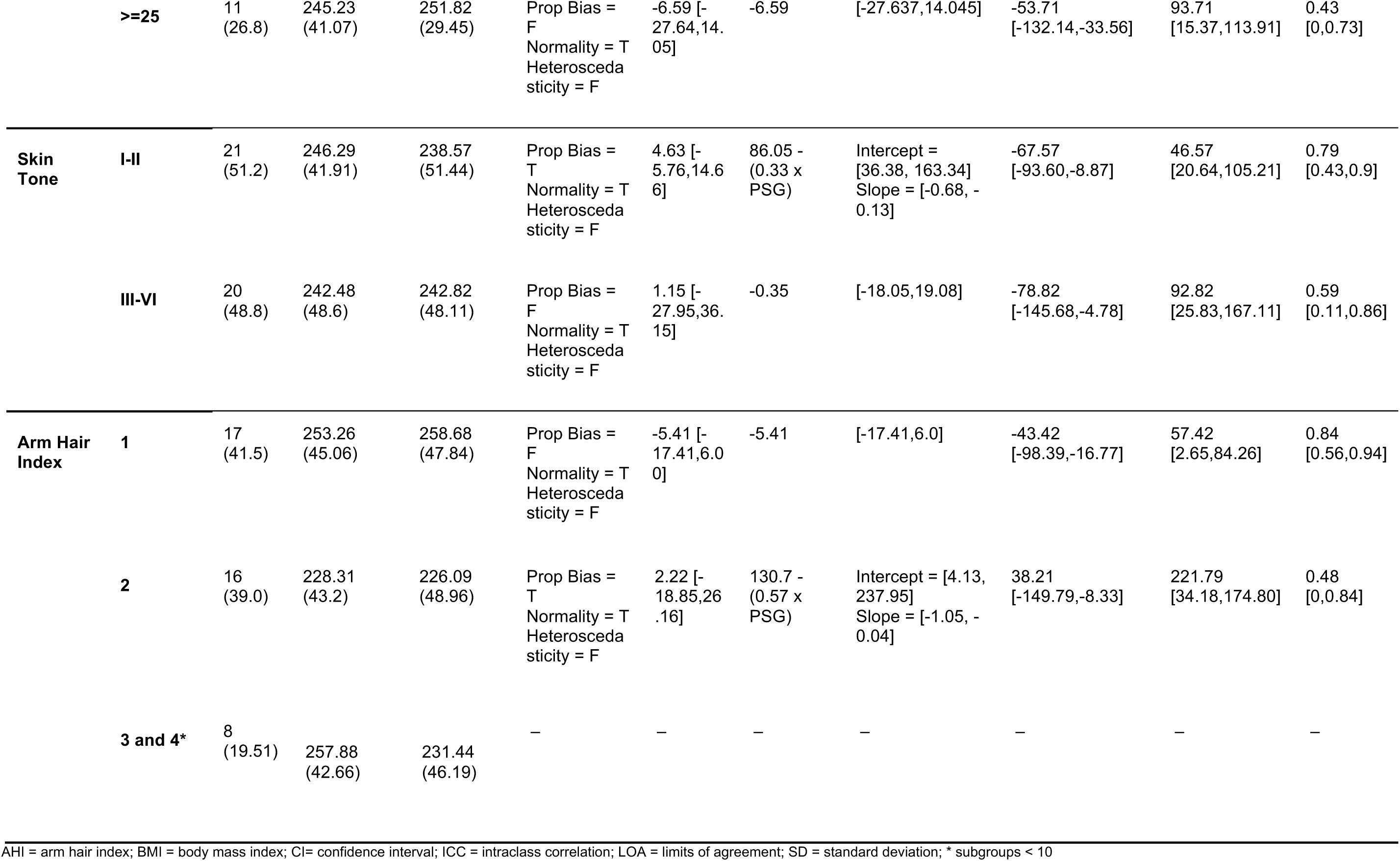
Summary of performance metrics for duration of light sleep according to subgroups.

**Supplement Table 10.**
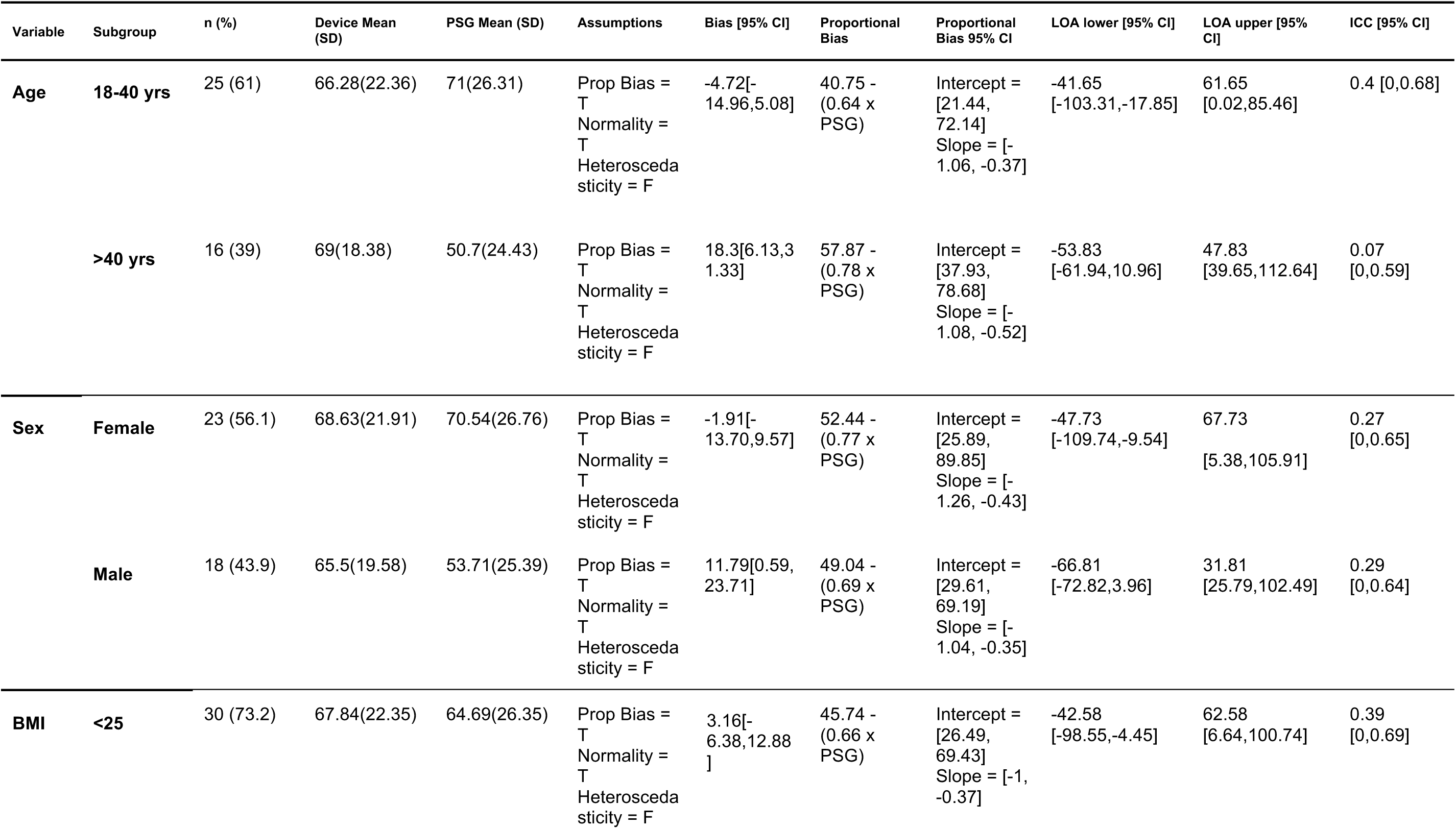
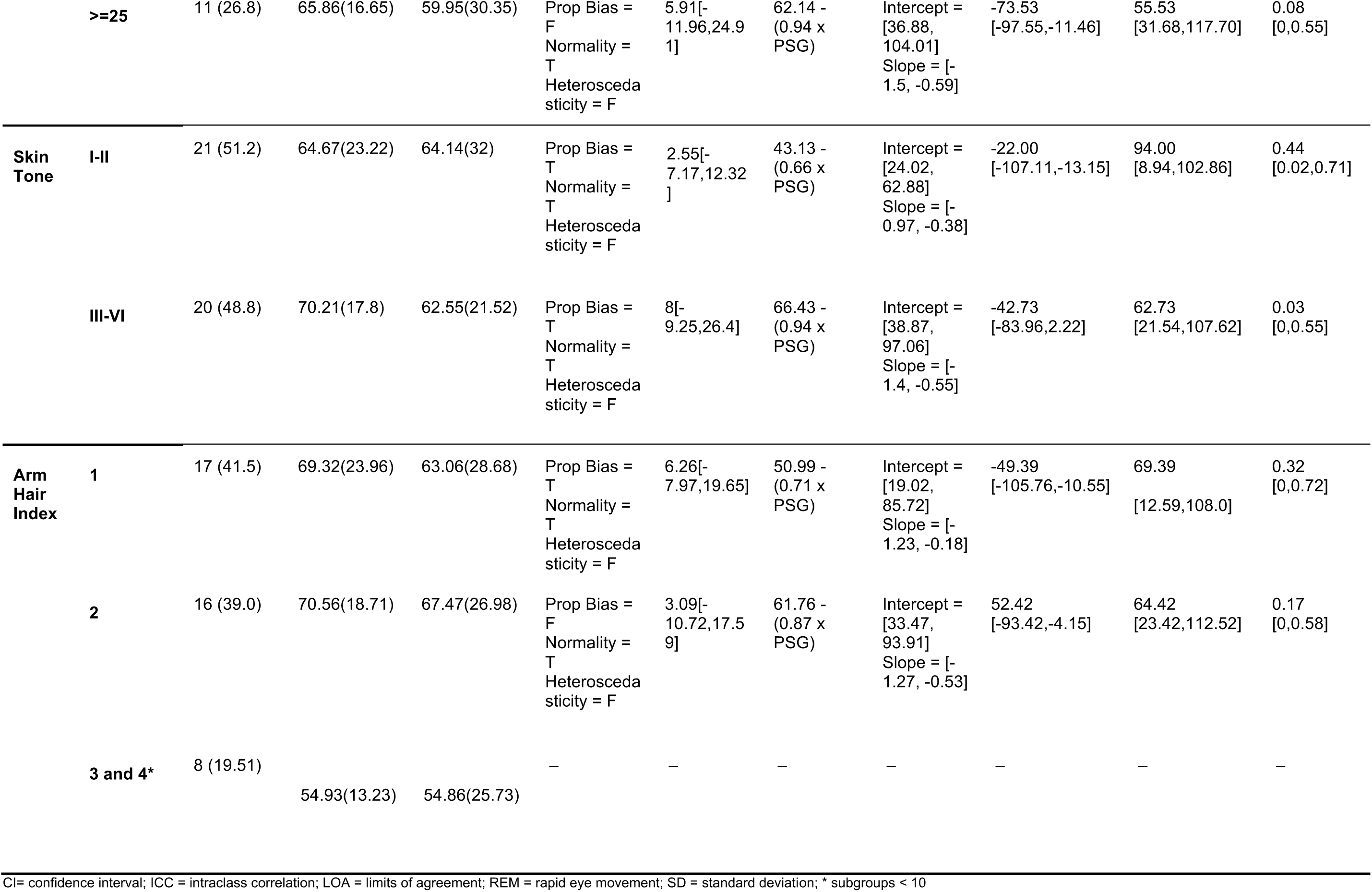
Summary of performance metrics for duration of deep sleep according to subgroups.

**Supplement Table 11.**
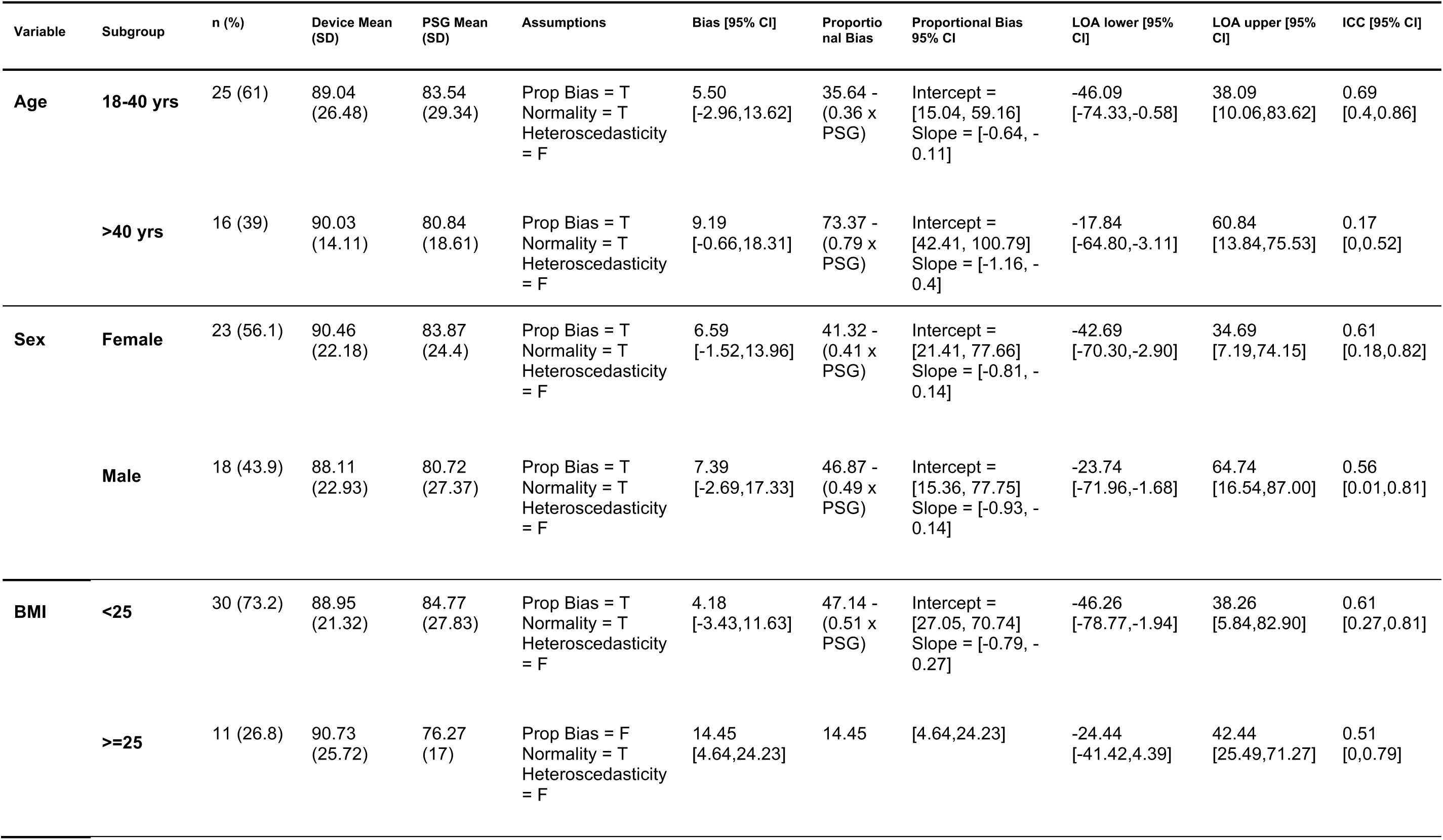
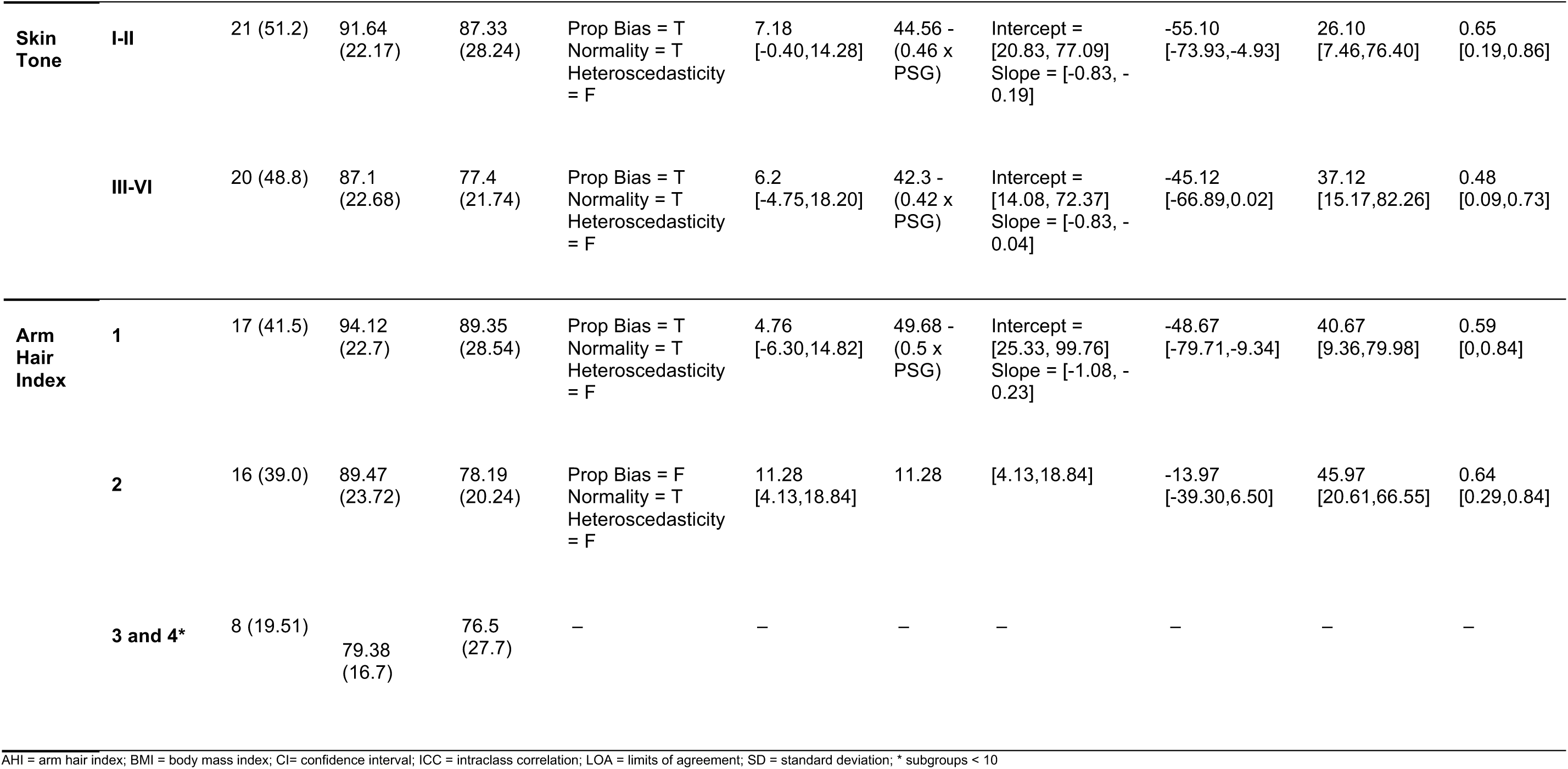
Summary of performance metrics for duration of REM sleep according to subgroups.

**Supplement Table 12.**
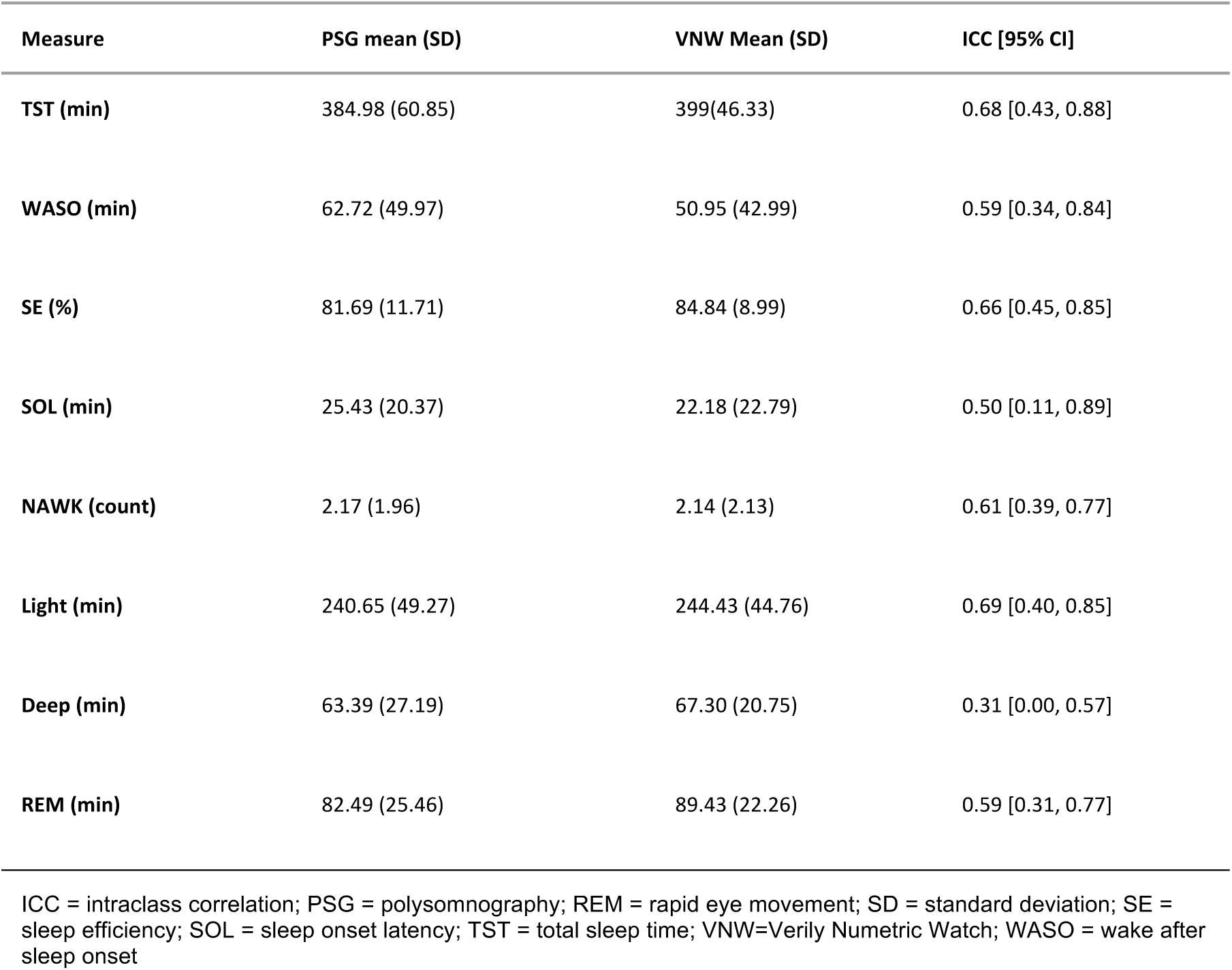
As an additional metric to evaluate the performance of the VNW algorithm, we calculated intra-class correlation coefficients between the mean values of each measure in both devices.

## Notes

### Competing Interest Statement

BWN, SSaeb, PB, NV, HA, NS, SSSullivan, SP, ER, RK, SShin report employment and equity ownership in Verily Life Sciences. In addition, SSaeb and NV are listed inventors in a pending patent broadly relevant to the work. MDZ, FB and NA received institutional research funding from Verily Life Sciences for study execution.

### Funding Statement

This study was funded by Verily Life Sciences

### Author Declarations

WCG Institutional Review Board (20215892) gave ethical approval for this work

