## Supplementary Materials for "Performance Evaluation of the Verily Numetric Watch sleep suite for digital sleep assessment against in-lab polysomnography"

Supplement Table 1.

Full summary of participant characteristics

| | | N=41 |
|---|---|---|
| Age (yrs) | Median (range) | 34.0 (18.0 - 78.0) |
| | Mean (SD) | 40.5 (16.5) |
| Age categories, n (%) | 18-40 | 25 (61.0) |
| | 40-60 | 8 (19.5) |
| | 60-80 | 8 (19.5) |
| Sex, n (%) | Female | 23 (56.1) |
| | Male | 18 (43.9) |
| BMI | Median (range) | 23.3 (17.8 - 36.0) |
| | Mean (SD) | 24.2 (4.2) |
| BMI categories, n (%) | <18.5 | 1 (2.4) |
| | 18.5-25 | 29 (70.7) |
| | 25-30 | 6 (14.6) |
| | ≥30 | 5 (12.2) |
| Ethnicity, n (%) | Hispanic or Latino | 4 (9.8) |
| | Not Hispanic or Latino | 37 (90.2) |
| Skin tone, n (%) | Type I - Always burns, never tans | 1 (2.44) |
| | Type II - Usually burns, then tans | 20 (48.8) |
| | Type III - May burn, tans well | 10 (24.4) |

| | | |
|---|---|---|
| | Type IV - Rarely burns, tans well | 5 (12.2) |
| | Type V - Very rarely burns, tans well, brown skin | 3 (7.3) |
| | Type VI - Very rarely burns, tans well, very dark skin | 2 (4.9) |
| Arm hair index, n (%) | 1: Little to no visible arm hair, light in color | 17 (41.5) |
| | 2: Visible, fine, arm hair, light to medium color | 16 (39.0) |
| | 3: Coarse arm hair, medium to dark color | 6 (14.6) |
| | 4: Very coarse arm hair, dark in color | 2 (4.9) |
| Right wrist circumference, cm | Median (range) | 6.2 (5.5 - 7.4) |
| | Mean (SD) | 6.29 (0.49) |
| Left wrist circumference, cm | Median (range) | 6.2 (5.3 - 7.5) |
| | Mean (SD) | 6.28 (0.48) |
| Right wrist circumference categories, n (%) | Large | 13 (31.7) |
| | Medium | 14 (34.1) |
| | Small | 14 (34.1) |
| Left wrist circumference categories, n (%) | Large | 13 (31.7) |
| | Medium | 14 (34.1) |
| | Small | 14 (34.1) |
| Race, n (%) | American Indian or Alaska Native | 1 (2.4) |
| | Asian | 8 (19.5) |
| | Black of African American | 4 (9.8) |
| | Mixed race | 4 (9.8) |

| | | |
|---|---|---|
| | Native Hawaiian or Other Pacific Islander | 1 (2.4) |
| | Other | 1 (2.4) |
| | White | 22 (53.7) |
| Dominant hand, n (%) | Ambidextrous | 1 (2.4) |
| | Left | 5 (12.2) |
| | Right | 35 (85.4) |
| OSA score | Median (range) | 0.0 (0.0 - 7.0) |
| | Mean (SD) | 1.34 (1.64) |
| ISI score | Median (range) | 3.0 (0.0 - 7.0) |
| | Mean (SD) | 2.95 (1.92) |
| ESS score | Median (range) | 5.0 (0.0 - 9.0) |
| | Mean (SD) | 5.0 (2.65) |
| AHI Index | Median (range) | 1.3 (0.1 - 4.7) |
| | Mean (SD) | 1.69 (1.2) |

AHI=Apnea hypopnea index ; BMI=body mass index; ESS=Epworth sleepiness scale ; ISI=insomnia severity index ; OSA=Obstructive Sleep Apnea; SD=standard deviation; Skin Type I and II were categorized as light skin tone; Skin Type III and IV were categorized as medium skin tone; Skin Type V and VI were categorized as dark skin tone

Detailed description of enrollment screening

Questionnaires completed by participants or study personnel and used for screening were:

- Obstructive Sleep Apnea 50 (OSA50): a brief 4-item (obesity, snoring, apneas, and age) diagnostic screening questionnaire for OSA in primary care that has been shown to identify patients with moderate to severe OSA with 84% accuracy [24]. This questionnaire has a cut-off score of ≥ 5, with typical sleepers having an OSA50 score of < 5 (sensitivity 100%, specificity 29%).
- AHI: calculated from PSG signals, it is the total number of apnea or hypopnea events in a night divided by the hours of sleep. The AHI assists in the diagnosis of OSA, classifying participants as having Normal sleep (< 5 events per hour), Mild (5-14 events per hour), Moderate (15-29 events per hour), and Severe (30 or more events per hour).
- Insomnia Severity Index (ISI): a 7-item questionnaire designed to screen for insomnia [25]. A 5-point Likert scale ranging from 0 = no problem to 4 = very severe problem is used for each question to calculate a total score ranging from 0 to 28. Total scores are categorized as absence of insomnia (0–7); sub-threshold insomnia (8–14); moderate insomnia (15–21); and severe insomnia (22–28). An ISI < 8 is used to identify no clinically significant insomnia symptoms.
- ESS: an 8-item self-reported questionnaire used to assess daytime sleepiness [30]. A 4-point Likert scale ranging from 0 = would never doze to 3 = high chance of dozing is used for each question to calculate a total score ranging from 0 to 24. Total scores are categorized as normal (0-10) and then increasing degree of daytime sleepiness (11-24). This measure has good internal consistency[43].
- Fitzpatrick Skin Scale (completed by study personnel) [31]: a 10-item questionnaire that assesses Genetic (physical traits), Sensitivity (reaction to sun exposure), and Intentional

Exposure (tanning habits) to categorize participants on a scale from 1-6 as research has shown that skin tone can influence the accuracy of PPG signals. Categories include, Type I (scores 0–6) always burns, never tans (palest; freckles), Type II (scores 7–13) usually burns, tans minimally (light colored but darker than fair), Type III (scores 14–20) sometimes mild burn, tans uniformly (golden honey or olive), Type IV (scores 21–27) burns minimally, always tans well (moderate brown), Type V (scores 28–34) very rarely burns, tans very easily (dark brown), Type VI (scores 35–36) never burns (deeply pigmented dark brown to darkest brown). These can be categorized into light (Type I and II), medium (Type III and IV), and dark (Type V and VI)[44].

- Arm Hair Index (scored by study personnel): assesses the density of participants' arm hair on a scale of one to four, including 1 (Little to no visible hair), 2 (Visible fine arm hair), 3 (Coarse arm hair), and 4 (Very coarse arm hair).

### Device synchronization during overnight study visit

Time synchronization between the VNW and PSG devices was performed using the following steps. [42]

First, the VNW was placed on the non-dominant wrist and the time (HH:MM) was recorded in the Device Accountability Log and electronic data capture (EDC) system.

Next, PSG hook up and calibration were performed; PSG was then turned on at the top of the minute on the VNW device (i.e., when the minute on the watch face changed) and the time (HH:MM) was logged in the EDC system.

Participants started wearing VNW before PSG was on and the overall difference between the onset times (mean = 3.51 seconds, SD = 4.06 seconds) was well below the range that has been shown to introduce significant bias in study outcomes. As is common practice, the start of the PSG recording was labeled as “Time 0” and time was rounded (e.g., 22:32:00).[42]

### Success Criteria for Sleep Versus Wake Classification as Product Requirement Specification

We pre-specified thresholds as “success criteria” for sleep versus wake classification with success being defined as sensitivity ≥ 0.90 and specificity ≥ 0.50 (the lower 95% CI bound over these values) as internal criteria for a successful sleep versus wake classification to proceed with subsequent development of the algorithm.

Supplement Table 2.

Summary of performance metrics for ‘sleep vs wake’ classification, according to participant subgroups

| Demographic Variable | Subgroup (n) | Sensitivity [95% CI] | Specificity [95% CI] | PPV [95% CI] | NPV [95% CI] |
|---|---|---|---|---|---|
| Sex | Female (23) | 0.95 [0.93, 0.98] | 0.66 [0.59, 0.75] | 0.93 [0.91, 0.96] | 0.73 [0.62, 0.88] |
| | Male (18) | 0.97 [0.97, 0.98] | 0.66 [0.59, 0.73] | 0.92 [0.89, 0.94] | 0.87 [0.82, 0.91] |
| Age | 18-40 (25) | 0.96 [0.95, 0.98] | 0.70 [0.63, 0.76] | 0.94 [0.92, 0.96] | 0.79 [0.70, 0.90] |
| | > 40 (16) | 0.96 [0.94, 0.98] | 0.62 [0.55, 0.71] | 0.90 [0.87, 0.94] | 0.79 [0.70, 0.91] |
| BMI | < 25 (30) | 0.96 [0.94, 0.98] | 0.65 [0.59, 0.72] | 0.92 [0.90, 0.94] | 0.76 [0.69, 0.85] |
| | 25 and greater (11) | 0.98 [0.97, 0.99] | 0.69 [0.60, 0.78] | 0.94 [0.93, 0.96] | 0.85 [0.80, 0.91] |
| Skin Tone | I - III (31) | 0.96 [0.95, 0.98] | 0.66 [0.62, 0.71] | 0.93 [0.91, 0.95] | 0.80 [0.72, 0.88] |
| | IV - VI (10) | 0.95 [0.92, 0.99] | 0.65 [0.52, 0.88] | 0.92 [0.86, 0.98] | 0.78 [0.65, 0.93] |
| Arm Hair Index | 1 (17) | 0.95 [0.92, 0.98] | 0.65 [0.58, 0.72] | 0.93 [0.91, 0.95] | 0.70 [0.57, 0.89] |
| | 2 (16) | 0.98 [0.97, 0.99] | 0.70 [0.61, 0.81] | 0.93 [0.91, 0.97] | 0.87 [0.81, 0.91] |
| | 3-4 (8)* | – | – | – | – |

BMI = body mass index; CI= confidence interval; NPV = negative predictive value; PPV = positive predictive value; * subgroups < 10

Supplement Table 3.

Summary of performance metrics for ‘sleep stage’’ classification, according to participant subgroups

| Variable | Subgroup (n) | Overall Cohen’s Kappa [95% CI] | Light Sleep Kappa [95% CI] | Deep Sleep Kappa [95% CI] | REM Sleep Kappa [95% CI] | Wake Kappa [95% CI] |
|---|---|---|---|---|---|---|
| Sex | Female (23) | 0.64[0.63,0.65] | 0.58[0.17,0.76] | 0.65[0.07,0.88] | 0.71[0.32,0.87] | 0.67[0.24,0.89] |
| | Male (18) | 0.69[0.68,0.70] | 0.62[0.46,0.77] | 0.67[0.31,0.87] | 0.75[0.56,0.91] | 0.67[0.45,0.87] |
| Age | 18-40 (25) | 0.68[0.67,0.69] | 0.63[0.41,0.78] | 0.7[0.24,0.89] | 0.72[0.48,0.87] | 0.69[0.34,0.89] |
| | > 40 (16) | 0.62[0.61,0.63] | 0.55[0.16,0.74] | 0.58[0.13,0.85] | 0.74[0.35,0.91] | 0.65[0.29,0.86] |
| BMI | < 25 (30) | 0.65[0.64,0.65] | 0.59[0.19,0.77] | 0.66[0.09,0.87] | 0.72[0.33,0.91] | 0.66[0.25,0.89] |
| | 25 and greater (11) | 0.69[0.68,0.7] | 0.62[0.47,0.76] | 0.66[0.31,0.87] | 0.76[0.65,0.87] | 0.71[0.47,0.87] |
| Skin Tone | I - III (31) | 0.67[0.66,0.68] | 0.61[0.4,0.78] | 0.63[0.12,0.87] | 0.74[0.48,0.91] | 0.67[0.34,0.84] |
| | IV - VI (10) | 0.65[0.64,0.66] | 0.59[0.18,0.76] | 0.69[0.24,0.87] | 0.72[0.37,0.89] | 0.67[0.3,0.9] |
| Arm Hair Index | 1 (17) | 0.62[0.61,0.63] | 0.57[0.19,0.77] | 0.59[0.04,0.87] | 0.69[0.3,0.85] | 0.64[0.25,0.81] |
| | 2 (16) | 0.7[0.69,0.71] | 0.63[0.38,0.76] | 0.73[0.39,0.88] | 0.76[0.6,0.91] | 0.73[0.45,0.91] |
| | 3-4 (8)* | – | – | – | – | – |

AHI = arm hair index; BMI = body mass index; CI= confidence interval; REM = rapid eye movement; * subgroups < 10

Supplement Table 4.

Summary of performance metrics for TST according to subgroups.

| Variable | Subgroup | n (%) | Device Mean (SD) | PSG Mean (SD) | Assumptions | Bias [95% CI] | Proportional Bias | Proportional Bias 95% CI | LOA lower [95% CI] | LOA upper [95% CI] | ICC [95% CI] |
|---|---|---|---|---|---|---|---|---|---|---|---|
| **Age** | **18-40 yrs** | 25 (61) | 397.58 (54.19) | 388.3 (66.32) | Prop Bias = T Normality = F Heteroscedasticity = F | 9.28 [-7.56,23.82] | 146.6 - (0.35 x PSG) | Intercept = [23.02,253.59] Slope = [-0.65, -0.03] | -66.59 [-144.23,-6.18] | 92.59 [15.11,153.35] | 0.76 [0.49,0.94] |
| | **>40 yrs** | 16 (39) | 402.56(31.77) | 379.78 (52.81) | Prop Bias = T Normality = F Heteroscedasticity = F | 22.78 [5.062,45.16] | 266.64 - (0.64 x PSG) | Intercept = [94.25, 375.24] Slope = [-0.93, -0.2] | -92.71 [-107.27,19.59] | 73.71 [59.15,185.76] | 0.42 [0,0.82] |
| **Sex** | **Female** | 23 (56.1) | 407.52(51.21) | 400.11 (62.33) | Prop Bias = F Normality = F Heteroscedasticity = F | 7.41 [-11.305,26.17] | 7.41 | [-11.31,26.17] | -78.50 [-165.53,-8.4] | 104.50 [17.31,175.73] | 0.66 [0.26,0.95] |
| | **Male** | 18 (43.9) | 389.31(38.2) | 365.64 (54.59) | Prop Bias = T Normality = T Heteroscedasticity = F | 23.67 [9.778,38.972] | 181.11 - (0.43 x PSG) | Intercept = [93.36, 253.85] Slope = [-0.64, -0.2] | -58.37 [-75.85,18.20] | 68.37 [50.87,144.60] | 0.65 [0.31,0.86] |
| **BMI** | **<25** | 30 (73.2) | 398.68(46.77) | 383.85 (66.08) | Prop Bias = T Normality = F Heteroscedasticity = F | 14.83 [-1.867,31.38] | 207.78 - (0.5 x PSG) | Intercept = [82.6, 303.29] Slope = [-0.76, -0.18] | -79.16 [-157.73,14.65] | 105.16 [25.11,199.68] | 0.63 [0.34,0.88] |
| | **>=25** | 11 (26.8) | 401.82(47.25) | 388.05 (46.21) | Prop Bias = F Normality = T Heteroscedasticity = F | 13.77 [2.409,25.46] | 13.77 | [2.41,25.46] | -20.44 [-51.91,2.98] | 60.44 [29.07,83.79] | 0.86 [0.44,0.95] |

| | | | | | | | | | | | |
|---|---|---|---|---|---|---|---|---|---|---|---|
| **Skin Tone** | **I-II** | 21 (51.2) | 402.6 (49.94) | 390.05 (66.52) | Prop Bias = T Normality = F Heteroscedasticity = F | 14.29 [-0.647,26.44] | 183.25 - (0.44 x PSG) | Intercept = [52.05, 301.34] Slope = [-0.76, -0.1] | -75.39 [-156.01,-11.29] | 97.39 [16.90,161.99 ] | 0.7 [0.35,0.92] |
| | **III-VI** | 20 (48.8) | 396.3 (43.26) | 379.65 (55.49) | Prop Bias = T Normality = F Heteroscedasticity = F | 15.35 [-6.90,49.20] | 186.36 - (0.45 x PSG) | Intercept = [15.78, 321.85] Slope = [-0.8, -0.02] | -64.05 [-100.39,22.02] | 90.05 [53.60,175.66 ] | 0.64 [0.2,0.92] |
| **Arm Hair Index** | **1** | 17 (41.5) | 416.71(49.4) | 411.09 (57.12) | Prop Bias = T Normality = F Heteroscedasticity = F | 5.62 [-17.176,22.12] | 171.52 - (0.4 x PSG) | Intercept = [32.85, 385.14] Slope = [-0.93, -0.05] | -70.42 [-162.597,-36.82] | 96.42 [3.75,130.13] | 0.68 [0.07,0.94] |
| | **2** | 16 (39.0) | 388.34(45.21) | 371.75 (55.94) | Prop Bias = F Normality = F Heteroscedasticity = F | 16.59 [1.69,38.10] | 16.59 | [1.69,38.10] | 75.00 [-89.40,14.84] | 229.00 [64.59,169.47 ] | 0.65 [0.05,0.96] |
| | **3 and 4*** | 8 (19.51) | 385.38 (32.77) | 355.94 (63.05) | – | – | – | – | – | – | – |

AHI = arm hair index; BMI = body mass index; CI= confidence interval; ICC = intraclass correlation; LOA = limits of agreement; SD = standard deviation; TST = total sleep time; * subgroups < 10

Supplement Table 5.

Summary of performance metrics for WASO according to subgroups.

| Variable | Subgroup | n (%) | Device Mean (SD) | PSG Mean (SD) | Assumptions | Bias [95% CI] | Proportional Bias | Proportional Bias 95% CI | LOA lower [95% CI] | LOA upper [95% CI] | ICC [95% CI] |
|---|---|---|---|---|---|---|---|---|---|---|---|
| **Age** | **18-40 yrs** | 25 (61) | 44.68 (47.55) | 48.2 (44.41) | Prop Bias = T Normality = F Heteroscedasticity = F | -3.52 [-16.48, 13.60] | 11.76 - (0.32 x PSG) | Intercept = [0.07, 29.58] Slope = [-0.47, -0.04] | -82.91 [-128.63,-0.20] | 70.91 [25.05, 153.48] | 0.63 [0.32,0.89] |
| | **>40 yrs** | 16 (39) | 60.75 (33.81) | 85.41 (51.02) | Prop Bias = F Normality = F Heteroscedasticity = F | -24.66 [-47.06,-7.09] | -24.66 | [-47.06,-7.09] | -71.81 [-186.0,-63.26] | 91.81 [-22.82, 100.21] | 0.41 [0,0.83] |
| **Sex** | **Female** | 23 (56.1) | 49.11 (43.77) | 57.63 (49.57) | Prop Bias = T Normality = F Heteroscedasticity = F | -8.52 [-28.91, 12.24] | 27.95 - (0.63 x PSG) | Intercept = [4.05, 54.09] Slope = [-0.86, -0.14] | -105.34 [-194.29,-18.91] | 93.34 [3.81, 180.63] | 0.4 [0.13,0.83] |
| | **Male** | 18 (43.9) | 53.31 (43.13) | 69.22 (51.15) | Prop Bias = T Normality = T Heteroscedasticity = F | -15.92 [-27.17,-5.42] | 2.17 - (0.26 x PSG) | Intercept = [-11.33, 12.63], Slope = [-0.45, -0.01] | -55.42 [-102.84,-30.56] | 41.42 [-5.90, 66.27] | 0.81 [0.64,0.9] |
| **BMI** | **<25** | 30 (73.2) | 55.87 (46.02) | 68.58 (55.18) | Prop Bias = T Normality = F Heteroscedasticity = F | -12.72 [-28.62, 4.02] | 21.65 - (0.5 x PSG) | Intercept = [2.68, 45.33] Slope = [-0.74, -0.2] | -96.34 [-183.94,-22.25] | 84.34 [-3.85, 159.51] | 0.56 [0.28,0.86] |

| | | | | | | | | | | | |
|---|---|---|---|---|---|---|---|---|---|---|---|
| | **>=25** | 11 (26.8) | 37.55 (31.35) | 46.73 (27.85) | Prop Bias = F<br>Normality = T<br>Heteroscedasticity = T | -9.18 [-22.18, 4.41] | -9.18 | [-22.18,4.41] | bias - 2.46(0.72 + 0.33 x PSG)<br>Intercept = [-11.4, 12.84]<br>Slope = [0, 0.66] | bias + 2.46(0.72 + 0.33 x PSG<br>Intercept = [-11.4, 12.84]<br>Slope = [0, 0.66] | 0.63 [0,0.83] |
| **Skin Tone** | **I-II** | 21 (51.2) | 53.64 (49.59) | 60.10 (44.02) | Prop Bias = T<br>Normality = F<br>Heteroscedasticity = F | -7.55 [-18.92, 6.73] | 13.9 - (0.34 x PSG) | Intercept = [-6.89, 45.32]<br>Slope = [-0.64, -0.04] | -83.44 [-136.77,-4.91] | 84.44 [31.01,163.01] | 0.58 [0.21,0.88] |
| | **III-VI** | 20 (48.8) | 48.12 (35.86) | 65.47 (56.59) | Prop Bias = T<br>Normality = F<br>Heteroscedasticity = T | -24.85 [-58.4,-1.85] | 18.13 - (0.54 x PSG) | Intercept = [0.75, 33.01]<br>Slope = [-0.75, -0.15] | bias - 2.46(7.55 + 0.16 x PSG)<br>Intercept = [2.13, 14.65], Slope = [0.02, 0.27] | bias + 2.46(7.55 + 0.16 x PSG<br>Intercept = [2.13, 14.65]<br>Slope = [0.02, 0.27] | 0.6 [0.28,0.88] |
| **Arm Hair Index** | **1*** | 17 (41.5) | 50.88 (47.57) | 59.79 (43.54) | Prop Bias = T<br>Normality = F<br>Heteroscedasticity = F | -8.91[-26.118 ,15.35] | 18.8 - (0.46 x PSG) | Intercept = [-1.23, 57.06]<br>Slope = [-0.76, -0.26] | -96.33 [-142.43,-10.65] | 84.33<br>[38.12, 170.67] | 0.47 [0.1,0.84] |
| | **2*** | 16 (39.0) | 48.53 (43.85) | 58.38 (55.36) | Prop Bias = F<br>Normality = F<br>Heteroscedasticity = T | -9.84[-32.31, 5.85] | -9.84 | [-32.31,5.85] | bias - 2.46(2.37 + 0.34 x PSG)<br>Intercept = [-3.77, 9.5]<br>Slope = [0.08, 0.5] | bias + 2.46(2.37 + 0.34 x PSG)<br>Intercept = [-3.77, 9.5]<br>Slope = [0.08, 0.5] | 0.63 [0.28,0.95] |
| | **3 and 4*** | 8 (19.51) | 55.94 (35.04) | 77.62 (55.25) | – | – | – | – | – | – | – |

AHI = arm hair index; BMI = body mass index; CI= confidence interval; ICC = intraclass correlation; LOA = limits of agreement; SD = standard deviation; WASO = wake after sleep onset; * subgroups < 10

Supplement Table 6.

Summary of performance metrics for SE according to subgroups.

| Variable | Subgroup | n (%) | Device Mean (SD) | PSG Mean (SD) | Assumptions | Bias [95% CI] | Proportional Bias | Proportional Bias 95% CI | LOA lower [95% CI] | LOA upper [95% CI] | ICC [95% CI] |
|---|---|---|---|---|---|---|---|---|---|---|---|
| **Age** | **18-40 yrs** | 25 (61) | 85.65 (9.98) | 83.52 (11.91) | Prop Bias = T Normality = F Heteroscedasticity = F | 2.13 [-0.94,5.10] | 32.84 - (0.37 x PSG) | Intercept = [13.73, 49.15] Slope = [-0.56, -0.14] | -12.53 [-27.21,0.27] | 18.35 [3.61,31.15] | 0.73 [0.46,0.92] |
| | **>40 yrs** | 16 (39) | 83.58 (7.29) | 78.84 (11.16) | Prop Bias = T Normality = F Heteroscedasticity = T | 4.74 [1.13,9.04] | 50.05 - (0.57 x PSG) | Intercept = [7.85, 69] Slope = [-0.8, -0.07] | bias - 2.46(17.57 + -0.17 x PSG)<br><br>Intercept = [3.88, 32.5] Slope = [-0.35, -0.01] | bias + 2.46(17.57 + -0.17 x PSG) Intercept = [3.88, 32.5] Slope = [-0.35, -0.01] | 0.5 [0.15,0.84] |
| **Sex** | **Female** | 23 (56.1) | 85.18 (9.59) | 83.51 (11.14) | Prop Bias = F Normality = F Heteroscedasticity = F | 1.68 [-1.86,5.24] | 1.68 | [-1.86,5.24] | -14.43 [-30.52,-0.34] | 20.25 [4.11,34.21] | 0.63 [0.29,0.93] |
| | **Male** | 18 (43.9) | 84.41 (8.41) | 79.38 (12.33) | Prop Bias = T Normality = T Heteroscedasticity = F | 5.03 [2.04,8.18] | 38.52 - (0.42 x PSG) | Intercept = [21.41, 52.15] Slope = [-0.58, -0.22] | -12.40 [-16.19,3.63] | 14.45 [10.69,30.50] | 0.69 [0.46,0.86] |

| | | | | | | | | | | | |
|---|---|---|---|---|---|---|---|---|---|---|---|
| **BMI** | **<25** | 30 (73.2) | 83.99 (9.6) | 80.78 (13) | Prop Bias = T Normality = F Heteroscedasticity = F | 3.21 [0.03,6.49] | 41.55 - (0.47 x PSG) | Intercept = [19.13, 57.45] Slope = [-0.67, -0.21] | -15.01 [-29.86,4.03] | 20.83 [6.01,39.66] | 0.65 [0.41,0.86] |
| | **>=25** | 11 (26.8) | 87.17 (6.9) | 84.18 (7.02) | Prop Bias = F Normality = T Heteroscedasticity = F | 3.00 [0.52,5.63] | 3.00 | [0.52,5.63] | -4.78 [-11.48,0.87] | 13.12 [6.42,18.7] | 0.7 [0,0.92] |
| **Skin Tone** | **I-II** | 21 (51.2) | 84.90 (9.74) | 82.04 (11.95) | Prop Bias = T Normality = F Heteroscedasticity = F | 3.13 [0.42,5.68] | 37.54 - (0.42 x PSG) | Intercept = [18.54, 58.62] Slope = [-0.69, -0.19] | -14.66 [-29.27,-0.58] | 18.81 [4.09,32.92] | 0.66 [0.29,0.89] |
| | **III-VI** | 20 (48.8) | 84.79 (8.37) | 81.33 (11.76) | Prop Bias = F Normality = F Heteroscedasticity = F | 3.22 [-1.19,9.95] | 3.45 | [0.46,7.15 | -12.48 [-19.83,4.37] | 18.30 [10.98,35.31] | 0.65 [0.38,0.91] |
| **Arm Hair Index** | **1** | 17 (41.5) | 85.43 (10.67) | 83.95 (9.78) | Prop Bias = F Normality = F Heteroscedasticity = F | 1.48[ -2.56,4.57] | 1.48 | [-2.56,4.57] | -12.44 [-29.52,-5.81] | 18.25 [1.28,24.90] | 0.7 [0.22,0.92] |
| | **2** | 16 (39.0) | 84.91 (8.21) | 81.56 (12.6) | Prop Bias = T Normality = F Heterosce | 3.34 [0.37,7.59] | 42.7 - (0.48 x PSG) | Intercept = [7.99, 63.87] Slope = [-0.72, - | bias - 2.46(20.08 + -0.2 x PSG) Intercept = [11.08, 24.8] Slope = [-0.26, -0.1] | bias + 2.46(20.08 + -0.2 x PSG) Intercept = [11.08, 24.8] | 0.68 [0.37,0.95] |

| | | | | | | | | | | |
|---|---|---|---|---|---|---|---|---|---|---|
| | | | | dasticity = T | | | 0.08] | | Slope = [-0.26, -0.1] | |
| **3 and 4*** | 8 (19.51) | 83.48 (7.32) | 77.16 (13.73) | – | – | – | – | – | – | – |

AHI = arm hair index; BMI = body mass index; CI= confidence interval; ICC = intraclass correlation; LOA = limits of agreement; SD = standard deviation; SE = sleep efficiency; * subgroups < 10

Supplement Table 7.

Summary of performance metrics for SOL according to subgroups.

| Variable | Subgroup | n (%) | Device Mean (SD) | PSG Mean (SD) | Assumptions | Bias [95% CI] | Proportional Bias | Proportional Bias 95% CI | LOA lower [95% CI] | LOA upper [95% CI] | ICC [95% CI] |
|---|---|---|---|---|---|---|---|---|---|---|---|
| **Age** | **18-40 yrs** | 25 (61) | 23.3 (20.48) | 29.52 (22.07) | Prop Bias = F Normality = F Heteroscedasticity = T | -6.22 [-11.48,-2.02] | -6.22 | [-11.48,-2.02] | bias - 2.46(-0.09 + 0.27 x PSG) Intercept = [-3.28, 2.54], Slope = [0.15, 0.41] | bias + 2.46(-0.09 + 0.27 x PSG Intercept = [-3.28, 2.54], Slope = [0.15, 0.41] | 0.78 [0.44,0.97] |
| | **>40 yrs** | 16 (39) | 20.44 (26.62) | 19.03 (15.98) | Prop Bias = F Normality = F Heteroscedasticity = F | 1.41 [-11.56,17.38] | -3.59 | [-23.81,21.06] | -119.60 [-147.86,-12.57] | 63.60 [35.26,170.26] | 0.04 [0,0.89] |
| **Sex** | **Female** | 23 (56.1) | 23.87 (24.73) | 23.24 (17.79) | Prop Bias = T Normality = F Heteroscedasticity = F | 0.63 [-7.46,11.24] | 10.1-0.41 x PSG | Intercept = [-2.25, 33.15] Slope = [-1.12, -0.02] | -53.60 [-73.00,0.94] | 38.60 [19.44,93.00] | 0.4 [0,0.95] |
| | **Male** | 18 (43.9) | 20.03 (20.53) | 28.22 (23.5) | Prop Bias = F Normality = F Heteroscedasticity = T | -8.19 [-16.86,-1.31] | -8.19 | [-16.861,-1.306] | bias - 2.46(-0.62 + 0.38 x PSG) Intercept = [-4.23, 2.17] Slope = [0.24, 0.56] | bias + 2.46(-0.62 + 0.38 x PSG Intercept = [-4.23, 2.17] Slope = [0.24, 0.56] | 0.62 [0,0.97] |

| | | | | | | | | | | | |
|---|---|---|---|---|---|---|---|---|---|---|---|
| **BMI** | **<25** | 30 (73.2) | 22.48 (22.79) | 25.05 (19.3) | Prop Bias = T Normality = F Heteroscedasticity = F | -2.57 [-10.32,6.32] | 11.45 + -0.56 x PSG | Intercept = [-1.18, 28.35] Slope = [-1.06, -0.06] | -54.04 [-88.27,-6.33] | 39.04 [4.47,87.29] | 0.36 [0,0.89] |
| | **>=25** | 11 (26.8) | 21.36 (23.86) | 26.45 (24.04) | Prop Bias = F Normality = F Heteroscedasticity = T | -5.09 [-13.82,0.59] | -5.09 | [-13.82,0.59] | bias - 2.46(0.18 + 0.29 x PSG) Intercept = [-5.78, 3.39] Slope = [0.14, 0.7] | bias + 2.46(0.18 + 0.29 x PSG Intercept = [-5.78, 3.39] Slope = [0.14, 0.7] | 0.82 [0,0.99] |
| **Skin Tone** | **I-II** | 21 (51.2) | 19.71 (19.94) | 26.29 (23.49) | Prop Bias = T Normality = F Heteroscedasticity = T | -7.21 [-13.29,-2.26] | 4.08 + -0.41 x PSG | Intercept = [-2.53, 12.3] Slope = [-0.92, -0.01] | bias - 2.46(0.45 + 0.35 x PSG) Intercept = [-2.71, 2.97] Slope = [0.22, 0.53] | bias + 2.46(0.45 + 0.35 x PSG Intercept = [-2.71, 2.97] Slope = [0.22, 0.53] | 0.65 [0.03,0.96] |
| | **III-VI** | 20 (48.8) | 24.77 (25.71) | 24.52 (17.06) | Prop Bias = F Normality = F Heteroscedasticity = F | 9.05 [-1.90,29.05] | 0.25 | [-8.6,12.45] | -56.63 [-76.957, -2.9] | 41.63 [21.15,95.77] | 0.34 [0,0.96] |
| **Arm Hair Index** | **1** | 17 (41.5) | 23.03 (27.84) | 20.21 (16.92) | Prop Bias = F Normality = F Heteroscedasticity = F | 2.82 [-5.82,15.77] | 2.82 | [-5.82,15.765] | -57.12 [-64.47,2.21] | 42.12 [34.737,101.8] | 0.39 [0,0.95] |
| | **2** | 16 (39.0) | 21.88 (18.5) | 29.09 (20.46) | Prop Bias = F Normality = F Heteroscedasticity = | -7.22 [-17.13,0.03] | -7.22 | [-17.13,0.03] | bias - 2.46(-1.98 + 0.45 x PSG) Intercept = [-7.99, 2.6] Slope = [0.24, 0.61] | bias + 2.46(-1.98 + 0.45 x PSG Intercept = [-7.99, 2.6] Slope = [0.24, | 0.49 [0,0.99] |

| | | | | | | | | | | |
|---|---|---|---|---|---|---|---|---|---|---|
| | | | | T | | | | | 0.61] | |
| **3 and 4*** | 8 (19.51) | 21(21.42) | 29.19(26.58) | – | – | – | – | – | – | – |

AHI = arm hair index; BMI = body mass index; CI= confidence interval; ICC = intraclass correlation; LOA = limits of agreement; SD = standard deviation; SOL = sleep onset latency; * subgroups < 10

Supplement Table 8.

Summary of performance metrics for NAWK according to subgroups.

| Variable | Subgroup | n (%) | Device Mean (SD) | PSG Mean (SD) | Mean Difference [95%CI] | Kappa [95% CI] | ICC [95% CI] |
|---|---|---|---|---|---|---|---|
| **Age** | **18-40 yrs** | 25 (61) | 1.48(1.96) | 1.28(1.54) | 0.2[-0.52,0.96] | 0.29[0.09,0.52] | 0.43[0.13,0.7] |
| | **>40 yrs** | 16 (39) | 2.81(1.91) | 2.69(1.89) | 0.12[-0.56,0.81] | 0.49[0.17,0.66] | 0.7[0.44,0.84] |
| **Sex** | **Female** | 23 (56.1) | 2(1.76) | 1.74(1.71) | 0.26[-0.39,0.96] | 0.41[0.21,0.61] | 0.55[0.23,0.78] |
| | **Male** | 18 (43.9) | 2(2.38) | 1.94(1.95) | 0.06[-0.72,0.89] | 0.44[0.09,0.65] | 0.65[0.25,0.85] |
| **BMI** | **<25** | 30 (73.2) | 2.4(2.08) | 1.93(1.96) | 0.47[-0.13,1.13] | 0.39[0.22,0.58] | 0.58[0.29,0.78] |
| | **BMI >=25** | 11 (26.8) | 0.91(1.45) | 1.55(1.29) | -0.64[-1.18,-0.09] | 0.47[0.05,0.79] | 0.62[-0.27,0.87] |
| **Skin Tone** | **I-II** | 21 (51.2) | 2(2.1) | 1.62(1.6) | 0.38[-0.38,1.14] | 0.36[0.09,0.61] | 0.49[0.07,0.77] |
| | **III-VI** | 20 (48.8) | 2(2) | 2.05(2.01) | -0.05[-0.7,0.65] | 0.56[0.3,0.73] | 0.71[0.46,0.87] |

| | | | | | | | |
|---|---|---|---|---|---|---|---|
| **Arm Hair Index** | **1** | 17 (41.5) | 2.24(1.95) | 2(1.87) | 0.24[-0.53,1.12] | 0.39[0.12,0.62] | 0.57[0.15,0.84] |
| | **2** | 16 (39.0) | 1.81(2.26) | 1.5(1.79) | 0.31[-0.38,1.06] | 0.56[0.36,0.81] | 0.7[0.42,0.89] |
| | **3 and 4*** | 8 (19.51) | 1.88(1.89) | 2.12(1.81) | – | – | – |

CI= confidence interval; ICC = intraclass correlation; LOA = limits of agreement; NAWK = number of awakenings; SD = standard deviation; * subgroups < 10

Supplement Table 9.

Summary of performance metrics for duration of light sleep according to subgroups.

| Variable | Subgroup | n (%) | Device Mean (SD) | PSG Mean (SD) | Assumptions | Bias [95% CI] | Proportional Bias | Proportional Bias 95% CI | LOA lower [95% CI] | LOA upper [95% CI] | ICC [95% CI] |
|---|---|---|---|---|---|---|---|---|---|---|---|
| **Age** | **18-40 yrs** | 25 (61) | 242.26 (45.22) | 233.76 (49.14) | Prop Bias = T Normality = F Heteroscedasticity = F | 8.50 [-2.60,19.52] | 66.42 - (0.25 x PSG) | Intercept = [20.15, 127.66] Slope = [-0.52, -0.06] | -49.38 [-94.59,-7.81] | 63.38 [18.07,104.89] | 0.8 [0.56,0.91] |
| | **>40 yrs** | 16 (39) | 247.81 (45.29) | 251.41 (49.06) | Prop Bias = F Normality = F Heteroscedasticity = F | -3.59 [-23.81,21.06] | -3.59 | [-23.81,21.06] | -119.60 [-147.86,-12.57] | 63.60 [35.26,170.26] | 0.51 [0,0.83] |
| **Sex** | **Female** | 23 (56.1) | 248.43 (48.06) | 245.7 (57.12) | Prop Bias = T Normality = T Heteroscedasticity = F | 2.74 [-12.94,20.22] | 102.16 - (0.4 x PSG) | Intercept = [18.41, 186.98] Slope = [-0.76, -0.09] | -73.52 [-137.47,-0.31] | 87.52 [23.41,160.73] | 0.7 [0.32,0.89] |
| | **Male** | 18 (43.9) | 239.31 (40.94) | 234.19 (37.55) | Prop Bias = F Normality = T Heteroscedasticity = F | 5.11 [-9.44,19.11] | 5.11 | [-9.44,19.11] | -59.73 [-106.30,-13.76] | 63.73 [17.10,109.64] | 0.67 [0.11,0.86] |
| **BMI** | **<25** | 30 (73.2) | 244.13 (46.71) | 236.55 (54.63) | Prop Bias = T Normality = F Heteroscedasticity = F | 7.58 [-4.62,20.88] | 91.71 - (0.36 x PSG) | Intercept = [29.06, 167.53] Slope = [-0.68, -0.11] | -64.22 [-116.71,6.78] | 78.22 [25.88,149.09] | 0.73 [0.39,0.89] |

| | | | | | | | | | | | |
|---|---|---|---|---|---|---|---|---|---|---|---|
| | **>=25** | 11 (26.8) | 245.23 (41.07) | 251.82 (29.45) | Prop Bias = F<br>Normality = T<br>Heteroscedasticity = F | -6.59 [-27.64,14.05] | -6.59 | [-27.637,14.045] | -53.71 [-132.14,-33.56] | 93.71 [15.37,113.91] | 0.43 [0,0.73] |
| **Skin Tone** | **I-II** | 21 (51.2) | 246.29 (41.91) | 238.57 (51.44) | Prop Bias = T<br>Normality = T<br>Heteroscedasticity = F | 4.63 [-5.76,14.66] | 86.05 - (0.33 x PSG) | Intercept = [36.38, 163.34]<br>Slope = [-0.68, -0.13] | -67.57 [-93.60,-8.87] | 46.57 [20.64,105.21] | 0.79 [0.43,0.9] |
| | **III-VI** | 20 (48.8) | 242.48 (48.6) | 242.82 (48.11) | Prop Bias = F<br>Normality = T<br>Heteroscedasticity = F | 1.15 [-27.95,36.15] | -0.35 | [-18.05,19.08] | -78.82 [-145.68,-4.78] | 92.82 [25.83,167.11] | 0.59 [0.11,0.86] |
| **Arm Hair Index** | **1** | 17 (41.5) | 253.26 (45.06) | 258.68 (47.84) | Prop Bias = F<br>Normality = T<br>Heteroscedasticity = F | -5.41 [-17.41,6.00] | -5.41 | [-17.41,6.0] | -43.42 [-98.39,-16.77] | 57.42 [2.65,84.26] | 0.84 [0.56,0.94] |
| | **2** | 16 (39.0) | 228.31 (43.2) | 226.09 (48.96) | Prop Bias = T<br>Normality = T<br>Heteroscedasticity = F | 2.22 [-18.85,26.16] | 130.7 - (0.57 x PSG) | Intercept = [4.13, 237.95]<br>Slope = [-1.05, -0.04] | 38.21 [-149.79,-8.33] | 221.79 [34.18,174.80] | 0.48 [0,0.84] |
| | **3 and 4*** | 8 (19.51) | 257.88 (42.66) | 231.44 (46.19) | – | – | – | – | – | – | – |

AHI = arm hair index; BMI = body mass index; CI= confidence interval; ICC = intraclass correlation; LOA = limits of agreement; SD = standard deviation; * subgroups < 10

Supplement Table 10.

Summary of performance metrics for duration of deep sleep according to subgroups.

| Variable | Subgroup | n (%) | Device Mean (SD) | PSG Mean (SD) | Assumptions | Bias [95% CI] | Proportional Bias | Proportional Bias 95% CI | LOA lower [95% CI] | LOA upper [95% CI] | ICC [95% CI] |
|---|---|---|---|---|---|---|---|---|---|---|---|
| **Age** | **18-40 yrs** | 25 (61) | 66.28(22.36) | 71(26.31) | Prop Bias = T Normality = T Heteroscedasticity = F | -4.72[-14.96,5.08] | 40.75 - (0.64 x PSG) | Intercept = [21.44, 72.14] Slope = [-1.06, -0.37] | -41.65 [-103.31,-17.85] | 61.65 [0.02,85.46] | 0.4 [0,0.68] |
| | **>40 yrs** | 16 (39) | 69(18.38) | 50.7(24.43) | Prop Bias = T Normality = T Heteroscedasticity = F | 18.3[6.13,31.33] | 57.87 - (0.78 x PSG) | Intercept = [37.93, 78.68] Slope = [-1.08, -0.52] | -53.83 [-61.94,10.96] | 47.83 [39.65,112.64] | 0.07 [0,0.59] |
| **Sex** | **Female** | 23 (56.1) | 68.63(21.91) | 70.54(26.76) | Prop Bias = T Normality = T Heteroscedasticity = F | -1.91[-13.70,9.57] | 52.44 - (0.77 x PSG) | Intercept = [25.89, 89.85] Slope = [-1.26, -0.43] | -47.73 [-109.74,-9.54] | 67.73 [5.38,105.91] | 0.27 [0,0.65] |
| | **Male** | 18 (43.9) | 65.5(19.58) | 53.71(25.39) | Prop Bias = T Normality = T Heteroscedasticity = F | 11.79[0.59, 23.71] | 49.04 - (0.69 x PSG) | Intercept = [29.61, 69.19] Slope = [-1.04, -0.35] | -66.81 [-72.82,3.96] | 31.81 [25.79,102.49] | 0.29 [0,0.64] |
| **BMI** | **<25** | 30 (73.2) | 67.84(22.35) | 64.69(26.35) | Prop Bias = T Normality = T Heteroscedasticity = F | 3.16[-6.38,12.88] | 45.74 - (0.66 x PSG) | Intercept = [26.49, 69.43] Slope = [-1, -0.37] | -42.58 [-98.55,-4.45] | 62.58 [6.64,100.74] | 0.39 [0,0.69] |

| | | | | | | | | | | | |
|---|---|---|---|---|---|---|---|---|---|---|---|
| | **>=25** | 11 (26.8) | 65.86(16.65) | 59.95(30.35) | Prop Bias = F Normality = T Heteroscedasticity = F | 5.91[-11.96,24.91] | 62.14 - (0.94 x PSG) | Intercept = [36.88, 104.01] Slope = [-1.5, -0.59] | -73.53 [-97.55,-11.46] | 55.53 [31.68,117.70] | 0.08 [0,0.55] |
| **Skin Tone** | **I-II** | 21 (51.2) | 64.67(23.22) | 64.14(32) | Prop Bias = T Normality = T Heteroscedasticity = F | 2.55[-7.17,12.32] | 43.13 - (0.66 x PSG) | Intercept = [24.02, 62.88] Slope = [-0.97, -0.38] | -22.00 [-107.11,-13.15] | 94.00 [8.94,102.86] | 0.44 [0.02,0.71] |
| | **III-VI** | 20 (48.8) | 70.21(17.8) | 62.55(21.52) | Prop Bias = T Normality = T Heteroscedasticity = F | 8[-9.25,26.4] | 66.43 - (0.94 x PSG) | Intercept = [38.87, 97.06] Slope = [-1.4, -0.55] | -42.73 [-83.96,2.22] | 62.73 [21.54,107.62] | 0.03 [0,0.55] |
| **Arm Hair Index** | **1** | 17 (41.5) | 69.32(23.96) | 63.06(28.68) | Prop Bias = T Normality = T Heteroscedasticity = F | 6.26[-7.97,19.65] | 50.99 - (0.71 x PSG) | Intercept = [19.02, 85.72] Slope = [-1.23, -0.18] | -49.39 [-105.76,-10.55] | 69.39 [12.59,108.0] | 0.32 [0,0.72] |
| | **2** | 16 (39.0) | 70.56(18.71) | 67.47(26.98) | Prop Bias = F Normality = T Heteroscedasticity = F | 3.09[-10.72,17.59] | 61.76 - (0.87 x PSG) | Intercept = [33.47, 93.91] Slope = [-1.27, -0.53] | 52.42 [-93.42,-4.15] | 64.42 [23.42,112.52] | 0.17 [0,0.58] |
| | **3 and 4*** | 8 (19.51) | 54.93(13.23) | 54.86(25.73) | – | – | – | – | – | – | – |

CI= confidence interval; ICC = intraclass correlation; LOA = limits of agreement; REM = rapid eye movement; SD = standard deviation; * subgroups < 10

Supplement Table 11.

Summary of performance metrics for duration of REM sleep according to subgroups.

| Variable | Subgroup | n (%) | Device Mean (SD) | PSG Mean (SD) | Assumptions | Bias [95% CI] | Proportional Bias | Proportional Bias 95% CI | LOA lower [95% CI] | LOA upper [95% CI] | ICC [95% CI] |
|---|---|---|---|---|---|---|---|---|---|---|---|
| **Age** | **18-40 yrs** | 25 (61) | 89.04 (26.48) | 83.54 (29.34) | Prop Bias = T Normality = T Heteroscedasticity = F | 5.50 [-2.96,13.62] | 35.64 - (0.36 x PSG) | Intercept = [15.04, 59.16] Slope = [-0.64, -0.11] | -46.09 [-74.33,-0.58] | 38.09 [10.06,83.62] | 0.69 [0.4,0.86] |
| | **>40 yrs** | 16 (39) | 90.03 (14.11) | 80.84 (18.61) | Prop Bias = T Normality = T Heteroscedasticity = F | 9.19 [-0.66,18.31] | 73.37 - (0.79 x PSG) | Intercept = [42.41, 100.79] Slope = [-1.16, -0.4] | -17.84 [-64.80,-3.11] | 60.84 [13.84,75.53] | 0.17 [0,0.52] |
| **Sex** | **Female** | 23 (56.1) | 90.46 (22.18) | 83.87 (24.4) | Prop Bias = T Normality = T Heteroscedasticity = F | 6.59 [-1.52,13.96] | 41.32 - (0.41 x PSG) | Intercept = [21.41, 77.66] Slope = [-0.81, -0.14] | -42.69 [-70.30,-2.90] | 34.69 [7.19,74.15] | 0.61 [0.18,0.82] |
| | **Male** | 18 (43.9) | 88.11 (22.93) | 80.72 (27.37) | Prop Bias = T Normality = T Heteroscedasticity = F | 7.39 [-2.69,17.33] | 46.87 - (0.49 x PSG) | Intercept = [15.36, 77.75] Slope = [-0.93, -0.14] | -23.74 [-71.96,-1.68] | 64.74 [16.54,87.00] | 0.56 [0.01,0.81] |
| **BMI** | **<25** | 30 (73.2) | 88.95 (21.32) | 84.77 (27.83) | Prop Bias = T Normality = T Heteroscedasticity = F | 4.18 [-3.43,11.63] | 47.14 - (0.51 x PSG) | Intercept = [27.05, 70.74] Slope = [-0.79, -0.27] | -46.26 [-78.77,-1.94] | 38.26 [5.84,82.90] | 0.61 [0.27,0.81] |
| | **>=25** | 11 (26.8) | 90.73 (25.72) | 76.27 (17) | Prop Bias = F Normality = T Heteroscedasticity = F | 14.45 [4.64,24.23] | 14.45 | [4.64,24.23] | -24.44 [-41.42,4.39] | 42.44 [25.49,71.27] | 0.51 [0,0.79] |

| | | | | | | | | | | | |
|---|---|---|---|---|---|---|---|---|---|---|---|
| **Skin Tone** | **I-II** | 21 (51.2) | 91.64 (22.17) | 87.33 (28.24) | Prop Bias = T Normality = T Heteroscedasticity = F | 7.18 [-0.40,14.28] | 44.56 - (0.46 x PSG) | Intercept = [20.83, 77.09] Slope = [-0.83, -0.19] | -55.10 [-73.93,-4.93] | 26.10 [7.46,76.40] | 0.65 [0.19,0.86] |
| | **III-VI** | 20 (48.8) | 87.1 (22.68) | 77.4 (21.74) | Prop Bias = T Normality = T Heteroscedasticity = F | 6.2 [-4.75,18.20] | 42.3 - (0.42 x PSG) | Intercept = [14.08, 72.37] Slope = [-0.83, -0.04] | -45.12 [-66.89,0.02] | 37.12 [15.17,82.26] | 0.48 [0.09,0.73] |
| **Arm Hair Index** | **1** | 17 (41.5) | 94.12 (22.7) | 89.35 (28.54) | Prop Bias = T Normality = T Heteroscedasticity = F | 4.76 [-6.30,14.82] | 49.68 - (0.5 x PSG) | Intercept = [25.33, 99.76] Slope = [-1.08, -0.23] | -48.67 [-79.71,-9.34] | 40.67 [9.36,79.98] | 0.59 [0,0.84] |
| | **2** | 16 (39.0) | 89.47 (23.72) | 78.19 (20.24) | Prop Bias = F Normality = T Heteroscedasticity = F | 11.28 [4.13,18.84] | 11.28 | [4.13,18.84] | -13.97 [-39.30,6.50] | 45.97 [20.61,66.55] | 0.64 [0.29,0.84] |
| | **3 and 4*** | 8 (19.51) | 79.38 (16.7) | 76.5 (27.7) | – | – | – | – | – | – | – |

AHI = arm hair index; BMI = body mass index; CI= confidence interval; ICC = intraclass correlation; LOA = limits of agreement; SD = standard deviation; * subgroups < 10

Supplement Table 12.

As an additional metric to evaluate the performance of the VNW algorithm, we calculated intra-class correlation coefficients between the mean values of each measure in both devices.

| Measure | PSG mean (SD) | VNW Mean (SD) | ICC [95% CI] |
|---|---|---|---|
| TST (min) | 384.98 (60.85) | 399(46.33) | 0.68 [0.43, 0.88] |
| WASO (min) | 62.72 (49.97) | 50.95 (42.99) | 0.59 [0.34, 0.84] |
| SE (%) | 81.69 (11.71) | 84.84 (8.99) | 0.66 [0.45, 0.85] |
| SOL (min) | 25.43 (20.37) | 22.18 (22.79) | 0.50 [0.11, 0.89] |
| NAWK (count) | 2.17 (1.96) | 2.14 (2.13) | 0.61 [0.39, 0.77] |
| Light (min) | 240.65 (49.27) | 244.43 (44.76) | 0.69 [0.40, 0.85] |
| Deep (min) | 63.39 (27.19) | 67.30 (20.75) | 0.31 [0.00, 0.57] |
| REM (min) | 82.49 (25.46) | 89.43 (22.26) | 0.59 [0.31, 0.77] |

ICC = intraclass correlation; PSG = polysomnography; REM = rapid eye movement; SD = standard deviation; SE = sleep efficiency; SOL = sleep onset latency; TST = total sleep time; VNW=Verily Numetric Watch; WASO = wake after sleep onset
